# Genomic epidemiology and evolutionary dynamics of respiratory syncytial virus group B in Kilifi, Kenya, 2015-17

**DOI:** 10.1101/2020.03.08.20032920

**Authors:** Everlyn Kamau, James R. Otieno, Nickson Murunga, John W. Oketch, Joyce M. Ngoi, Zaydah R. de Laurent, Anthony Mwema, Joyce U. Nyiro, Charles N. Agoti, D. James Nokes

## Abstract

Respiratory syncytial virus (RSV) circulates worldwide and is a leading cause of acute respiratory illness in young children. There is paucity of genomic data from purposively sampled populations by which to investigate evolutionary dynamics and transmission patterns of RSV. Here we present an analysis of 295 RSV group B genomes from Kilifi, coastal Kenya, sampled from individuals seeking outpatient care in 9 health facilities across a defined geographical area (890 km^2^), over 2 RSV epidemics between 2015 and 2017. RSVB diversity was characterized by multiple viral introductions into the area and co-circulation of distinct genetic groups or clusters, which transmitted and diversified locally but with varying frequency. Bayesian analyses indicated a strong spatially and temporally structured viral population suggesting extensive within-epidemic virus transmission. Phylogeographic analysis provided a strong support for epidemiological linkage from one central health facility to other facilities. Increase in relative diversity paralleled increase in seasonal viral incidence. Importantly, we identified a cluster of viruses (n=91) that emerged in the 2016/17 epidemic, carrying distinct amino-acid signatures including a novel non-synonymous change (K68Q) in antigenic site Ø in the Fusion gene. A different non-synonymous change K68N was recently associated with escape from a potent neutralizing monoclonal antibody (MEDI8897). RSVB diversity was additionally marked by signature non-synonymous substitutions that were unique to particular genomic clusters, some of which were under diversifying selection. Our findings provide insights into recent evolutionary and epidemiologic behaviors of RSV group B, and highlight possible emergence of a novel antigenic variant, which has implications on current prophylactic development strategies.

## Introduction

Respiratory syncytial virus (RSV) is the most common cause of acute lower respiratory tract infection in children <5 years of age worldwide, with an estimated associated mortality of up to 199,000 deaths per year mostly in developing countries (1,2). RSV is also an important cause of community-acquired pneumonia among hospitalized adults of all ages (3). RSV is a member of the family *Pneumoviridae*, subfamily *Orthopneumovirus*, has an enveloped, non-segmented, single-stranded, negative sense RNA genome of approximately 15,000 nucleotides encoding 11 proteins: NS1, NS2, N, P, M, SH, G, F, M2-1, M2-2, and L (4). RSV clinical isolates are classified into two groups (RSVA and RSVB) based on antigenic and genetic variability (5). Distinct genotypes of RSV have been shown to circulate locally and globally (6-9) and strains may vary from location to location in any given season with viruses identified in one location being similar to those from vastly different geographic locations identified in different years (10), suggestive of rapid global transmission. The available therapeutic modalities for RSV are chiefly supportive, and prophylactic treatment with RSV-specific neutralizing antibody is effective in reducing RSV morbidity in infants, but its use is currently limited to high-risk populations in high-resource settings (11). There is no licensed RSV vaccine for routine use in immunization, however, prophylactic vaccine candidates and monoclonal antibodies (mAbs) are now in advanced clinical trials (12).

We have previously characterised the community dynamics of RSV in the coastal region of Kenya, using sequences of the highly variable G glycoprotein gene (encoding the attachment protein) for RSV groups A and B and using whole genomes of RSVA genotype ON1, almost exclusively from samples from pneumonia patients admitted to the Kilifi County Hospital (13-15). From these studies, RSV displays high genetic diversity of locally circulating strains, within and between consecutive epidemics. Furthermore, recurrent RSV epidemics in Kilifi are depicted by sequential replacement of genotypes, over the long term, and high turnover of variants within genotypes in the short term (13,14). In the study reported here, samples arise from a design aimed to limit temporal, age-related, illness severity, geographical, and health care access bias. Recruitment was carried throughout a study location, from representative health facilities, simultaneously, and of patients of any age with mild acute respiratory symptoms (16).

Phylodynamic and phylogeographic methods have been used increasingly to study molecular epidemiology and evolutionary dynamics of virus populations e.g. Ebola, Zika, and Influenza (17-21). However, despite the importance of RSV to pneumonia hospitalisation and mortality among children (1), equivalent genome-scale studies to examine RSV transmission patterns and evolutionary dynamics within a community setting, are still few and remain a major gap in our understanding of what viral factors shape epidemiologic and evolutionary processes RSV at the local level. While most molecular studies on RSV have focused on the G glycoprotein gene because of its high genetic diversity and utility as a phylogenetic marker, genome-wide genetic signatures in RSV genome additionally inform on diversity and the adaptive mechanisms of RSV viruses following introduction into communities (15).

Here, we applied a combination of molecular clock, coalescent and discrete diffusion phylogeographic models to whole genome sequences to measure genomic diversity and deduce spatial and temporal circulation of RSVB in rural Kilifi, coastal Kenya. We sought to characterize introductions, transmission and spread of RSVB from samples collected through outpatient surveillance of respiratory viruses, analogous to studying community epidemics. We present estimates of evolutionary parameters such as the genomic rate of evolution and relative genetic diversity and describe spatial and temporal clustering patterns of viruses within each epidemic as demonstrated by phylogenetically distinct clades. In particular, we determine possible selection-driven emergence of a novel RSVB variant carrying distinct amino acid signatures. Our study significantly increases the number of publicly available complete RSVB genomes, which will enable further studies of RSV evolution.

## Materials and methods

### Study site and patient recruitment

The annual RSV epidemics in Kilifi, coastal Kenya, are seasonal starting from November through May, with a peak around January and an average duration of 18 weeks (22). The study from which the samples arise and used in the present report was done from December 2015 to March 2018, a period covering three RSV seasonal epidemics (2015/16, 2016/17 and 2017/18), and was carried out within the Kilifi Health and Demographic Surveillance System (KHDSS) (23). The study was conducted to document the community-wide burden of respiratory virus infections. Nine out of the 24 public outpatient health facilities in KHDSS were purposively selected (**Figure 1(A)**) – Matsangoni (MAT), Ngerenya (NGE), Mtondia (MTO), Sokoke (SOK), Mavueni (MAV), Jaribuni (JAR), Chasimba (CHA), Pingilikani (PIN) and Junju (JUN) - to provide a broad representation across the geographical region, covering major road networks into the location and variation in population density (16). Participant recruitment and specimen collection was integrated within the routine patient care and led by a resident clinician or nurse as detailed in (16). Patients of any age presenting with one or more ARI symptoms of cough, sneezing, nasal congestion, difficulty breathing, or increased respiratory rate for age were eligible. Written individual informed consent was sought from adult patients and parents/guardians of patients below 18 years.

**Figure 1.**
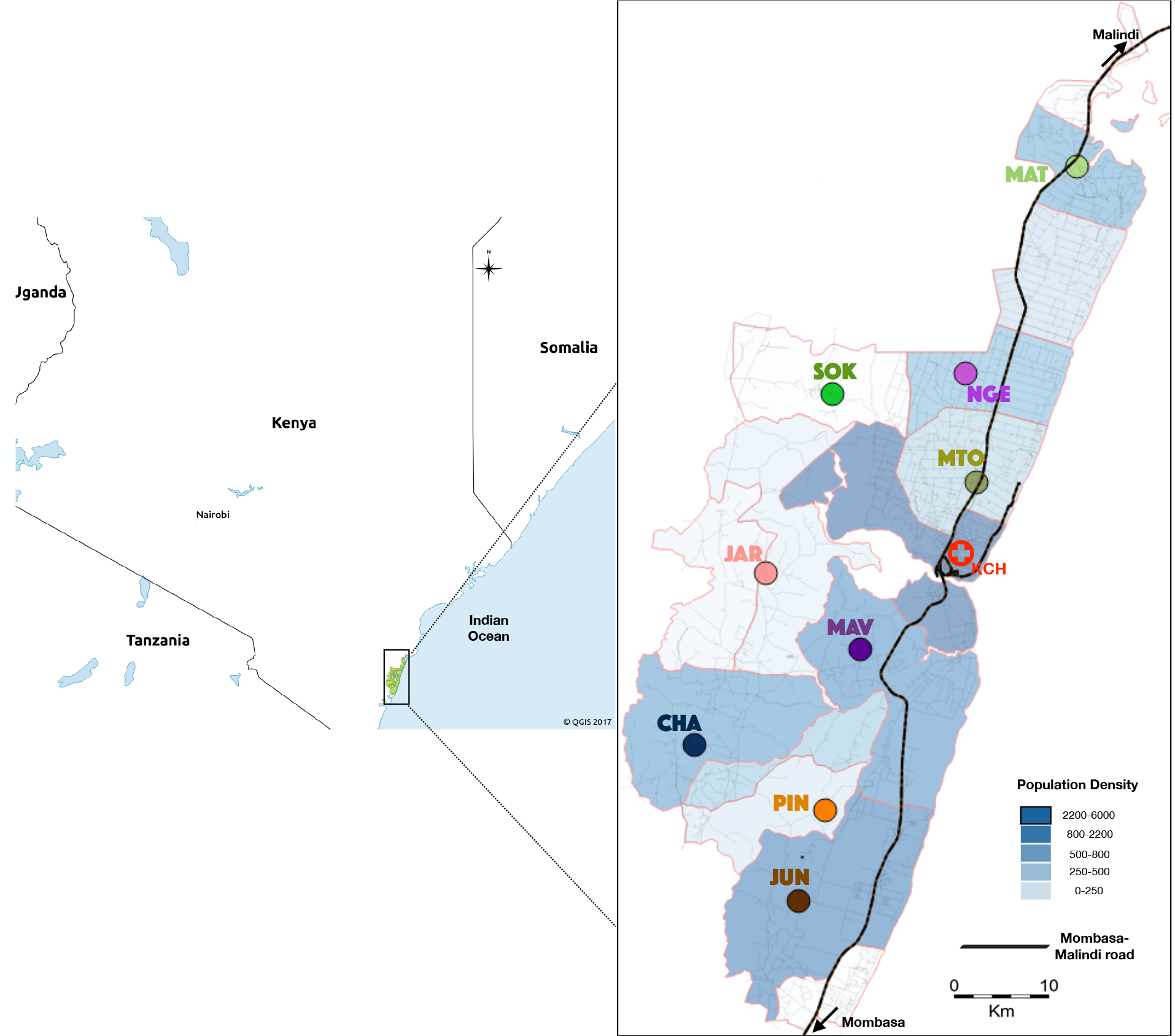
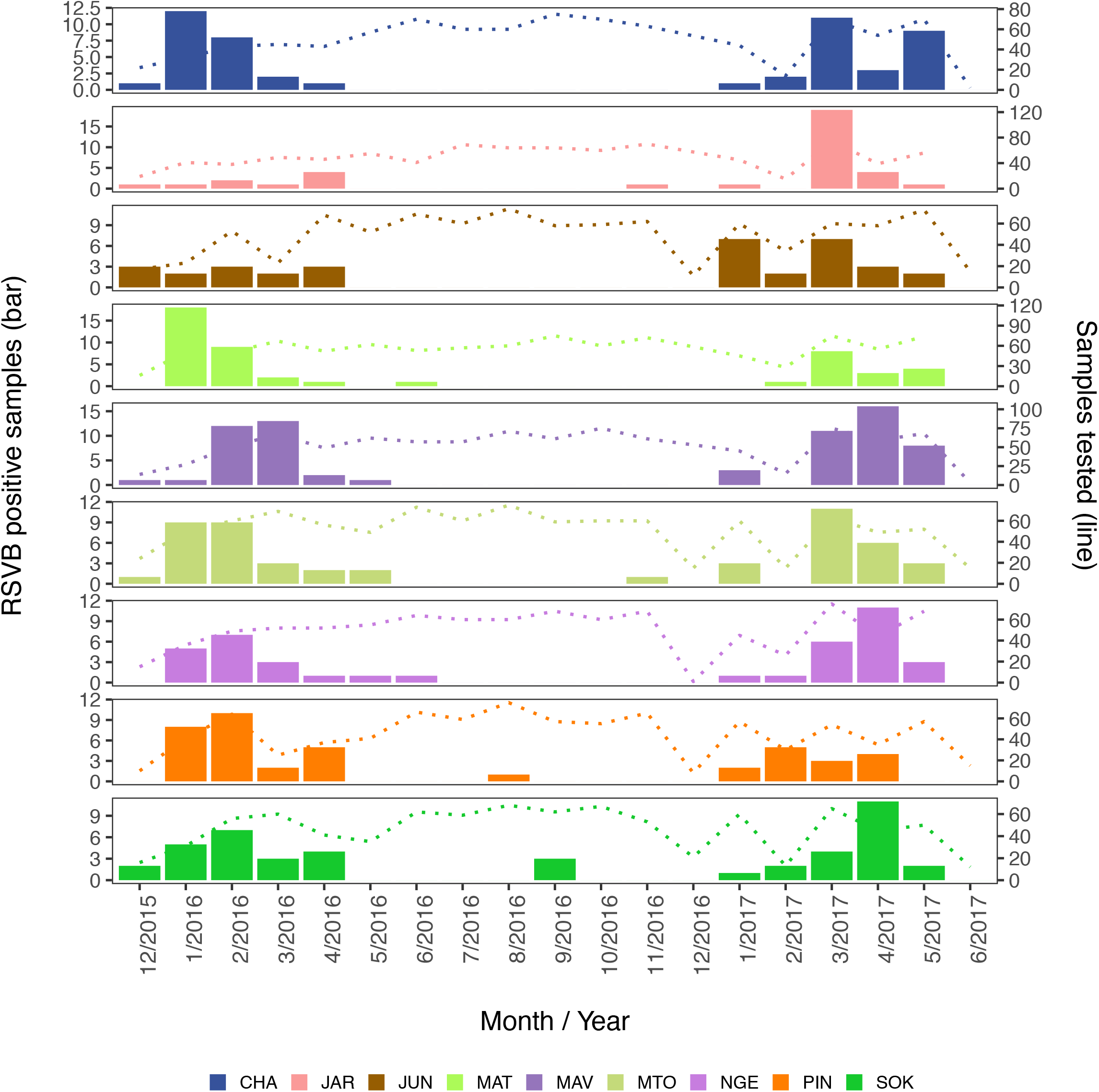
(**A**) A map showing the geographical area covered in the Kilifi Health Demographic Surveillance System (KHDSS), expanded from a map of Kenya. The nine participating public health facilities are indicated in the map. The dark lines within the polygons indicate the road structure within KHDSS. The maps were rendered using QGIS 2.18.17 (https://www.qgis.org/) (**B**) Monthly RSVB occurrence by study location: temporal and spatial distribution of RSVB positive cases (left Y axis) and number of clinical samples tested (right Y axis) from each participating health facilities. Abbreviations: CHA = Chasimba, JAR = Jaribuni, JUN = Junju, MAV = Mavueni, MAT = Matsangoni, PIN = Pingilikani, NGE = Ngerenya, SOK = Sokoke, MTO = Mtondia

The study was approved by the KEMRI-Scientific and Ethical Review Unit (SERU# 3103) and the University of Warwick Biomedical and Scientific Research Ethics Committee (BSREC# REGO-2015-6102).

### Sample collection and laboratory testing

A nasopharyngeal swab (NPS) was collected from each participant and stored in universal virus transport media (Copan Diagnostics, USA). RNA was extracted from samples by Qiacube HT using an RNeasy extraction kit (Qiagen, Germany) and screened for a range of respiratory viruses including RSVA and RSVB using a multiplex real-time PCR assay system (24,25). Samples with real time RT-PCR cycle threshold (Ct) of <35.0 were defined as positive for the target virus.

### RSVB whole genome amplification and sequencing

Whole genome amplification and sequencing was attempted for all RSVB positive samples with PCR cycle threshold (Ct) value < 35.0 collected between mid-December 2015 to end of May 2017. No RSVB was detected from June 2017 to the end of the study in March 2018 (i.e. 2017/18 epidemic was RSVA only). Reverse transcription of RNA molecules and PCR amplification were performed with a six-amplicon, six-reaction strategy presented in detail in (26). Briefly, extracted RNA was converted to cDNA using Superscript III First Strand Synthesis kit (Invitrogen) and forward primers, followed by PCR using Phusion High-Fidelity DNA polymerase (NEB). Amplification success was confirmed by observing the expected PCR product size (2300– 2500 bp) on 0.6% agarose gels. Amplicons were fragmented, tagged to adapters, and indexed using the Nextera XT (Illumina, San Diego, CA, USA) library prep kit as per manufacturer’s instructions. Size distribution of the barcoded libraries was assessed by Agilent’s 2100 Bioanalyzer. Pooled and normalized libraries in batches of seventy or eighty were sequenced on Illumina MiSeq system using 2⨯ 250 bp or 2 ⨯ 300 bp paired-end (PE) sequencing at the KEMRI-Wellcome Trust Research Program laboratories.

### Short read data assembly

Methods for quality check of the sequence reads, depletion of human reads, generation of consensus genome assemblies and calculation of coverage depth, were as described in (15). Briefly, quality check of the sequence reads was done using FastQC. Consensus genome assemblies were generated using viral-ngs pipeline v1.19.0 (Broad Institute). The raw reads were depleted of human reads by mapping onto the human reference genome hg19 using bowtie2 (27), and samtools (28) used to filter, sort, and recover the unmapped reads. The raw reads were mapped onto individual consensus assemblies with bowtie2, then samtools used to sort and index the aligned .bam files, and BEDtools (29) used to generate the coverage depth statistics.

### Sequence data compilation

To contextualize the diversity of RSVB in Kilifi in 2015 to 2017, the new Kilifi genomes were appended to a global dataset of other RSVB genomes available in Genbank (>14000 nucleotides (nt)), retrieved on 29 September 2019 (**Supplementary Table 1**). Only global sequences belonging to the RSVB BA genotype were considered, since the Kilifi sequences were of the BA genotype. The global dataset comprised sequences of viruses collected between 2012 and 2016 from UK, US, Russia, Philippines, Nicaragua, Jordan and India. Sequences of viruses sampled before 2012 and published without date or location of sampling were excluded.

### Phylogenetic and phylogeographic analyses

Sequence alignment was done using MAFFT v.7.221 (30) using the parameters ‘–localpair –maxiterate 1000’. To determine the degree of temporal signal of divergence in the Kilifi RSVB genomes, a maximum likelihood (ML) tree was estimated using RAxML (31) under general time-reversible (GTR) nucleotide substitution model with gamma-distributed among-site rate heterogeneity, and examined by exploratory linear regression analysis in Tempest v1.5.1 (32). jModelTest (33) was used to determine best-fit substitution model. Root-to-tip divergence was plotted as a function of sampling date (day-month-year). Branch support for ML trees was evaluated using 1000 bootstrap replicates.

Bayesian molecular clock phylogenies and discrete trait phylogeographic analyses were done using BEAST v1.10 (34) for 300 million MCMC steps, sampling parameters and trees every 5000 steps. HKY nucleotide substitution model with a gamma-distributed rate variation among sites was used. Path-sampling and stepping-stone marginal likelihood estimator (MLE) models (35) were used to estimate the most probable combination of uncorrelated lognormal relaxed molecular clock and coalescent models. The best fitting combination was the uncorrelated log-normal distributed relaxed molecular clock and Skygrid’s Gaussian Markov random field (GMRF) tree prior. A non-informative continuous time Markov chain reference prior was used on the molecular clock. Mixing and convergence of the MCMC sampler in the posterior target distribution was evaluated using Tracer v1.6 (http://beast.bio.ed.ac.uk/Tracer), and a maximum clade credibility (MCC) trees summarized with TreeAnnotator after removal of 10% burn-in. MCC trees were visualized with FigTree 1.4.4 (http://tree.bio.ed.ac.uk/software/figtree/).

To describe and quantify viral movement in Kilifi, we performed a discrete trait diffusion analysis, using sampling location as a discrete trait. We used a Bayesian stochastic search variable selection (BSSVS) procedure (36) and compared reversible and non-reversible (asymmetric) discrete diffusion phylogeographic models to estimate the most relevant transition pathways within the study locations. The standard implementation of the discrete asymmetrical phylogeographic approach assigns the same prior probability distribution to all of the transition rates in the continuous time markov chain (CTMC) (37). On the other hand, the reversible CTMC model allows more balanced transitions in the phylogeny or free lineage exchange between location states without preconditioned directionality but may have a poor fit for locations that could have unidirectional links in reality e.g. spatially expanding epidemics (36). Statistical support for migratory events was measured using Bayes factors (BF) and summarized using SpreaD3 software (38) after discarding 10% burn-in. We assumed that a BF > 6 is strong evidence for a well-supported viral pathway between two locations.

### Molecular evolution and adaptation

Gene-specific ratio of nonsynonymous to synonymous substitutions per site (dN/dS) were estimated for codon-based alignments using the single likelihood ancestor counting (SLAC) method available at the Datamonkey webserver (39). We also investigated episodic positive or diversifying selection using mixed-effects model of evolution (MEME) and Fast, Unconstrained Bayesian AppRoximation (FUBAR) approaches also available in Datamonkey. MEME aims to detect sites evolving under positive selection in a proportion of branches, while FUBAR uses a Bayesian approach and assumes that the selection pressure is constant along the entire phylogeny.

### Phylogenomic clusters

We defined a phylogenomic cluster as genetically linked viral sequences (≥2) that were more closely related than any randomly selected sequences within genetic distance threshold of *<d*. Maximum likelihood evolutionary distances, an estimate of the average number of changes per base pair, were calculated for each pair of consensus genomes using IQ-TREE (40) to measure genetic similarity or dissimilarity between pairs of taxa. ML distances, depicting genetic distance between sequences in a phylogenetic tree measured in nucleotide substitutions per site, were selected over the conventional nucleotide *p*-distances in order to correct for multiple nucleotide substitutions.

The distribution histogram plots of pairwise genetic distances were generated in R, and a distance cutoff value was determined as the least frequent value (*d*) between the first and second peaks in the histogram. Median values of pairwise ML evolutionary distance distributions of the identified phylogenomic cluster were checked if they were below *t*-percentile of the overall distance distribution. The *t*-percentile corresponded to the *d* distance threshold. In addition, a phylogenomic cluster was only considered eligible if it had reliability of ≥ 90% phylogenetic bootstrap support or Bayesian posterior probability. Each inferred phylogenomic cluster was evaluated for size, temporal and genetic characteristics.

### Phylogeny-trait association analysis

We used the Bayesian Tip-association Significance (BaTS) software (41), which uses the posterior sets of trees from Bayesian MCMC analysis, to measure the degree of association between sampling location and the phylogeny and are inversely correlated with the degree of association. Each sequence was assigned a location trait reflecting its origin of sampling and the first 10% of tree states were removed as burn-in. The overall statistical significance was determined by estimating the parsimony score (PS) and association index (AI) metrics, where the null hypothesis was that clustering by location was not more than expected by chance alone. In addition, the maximum clade size (MC) metric was used to compare the strength of clustering at each location by calculating the expected (null) and the observed mean clade size from each study location. A significance level of 0.05 was used in all cases. The PS, AI and MC statistics were computed for a null distribution with 1000 replicates.

### Sequence data availability

The final sequencing reads are available in the NCBI BioProject database under the study accession PRJNA562116 and the genomes generated in this study are available in GenBank under accession numbers MN365302 to MN365600.

## Results

### RSV occurrence in Kilifi, 2015 to 2017

Between December 2015 and June 2017, a total of 8127 nasopharyngeal swab samples were tested for RSV. RSV was detected in 503 (6.19%) samples (Ct <35): 95 (18.9%) were RSV group A (RSVA) and 408 (81.1%) were RSV group B (RSVB). The frequency and pattern of occurrence of RSVB for each health facility are shown in **Figure 1(B)**. The mean (SD) Ct value was 26.9 (4.16) and 25.0 (4.16) for RSVA and RSVB, respectively. The proportion of RSV positive individuals differed by age (*p* value <0.001), study location (*p* value = 0.003) but not by gender (*p* value = 0.078) (**Table 1**). The distribution of RSV viral load (equated to rRT-PCR Ct value) did not differ by outpatient health facility (**Supplementary Figure 1(A)**). The median age of RSV positive individuals was 20 months (interquartile range (IQR), 8-43 months), 81.7% (411/503) were aged 5 years or younger, and 272 (54.1%) of the cases were female (**Table 1**). RSV prevalence across different age groups by gender is shown in **Supplementary Figure 2**.

**Table 1.**
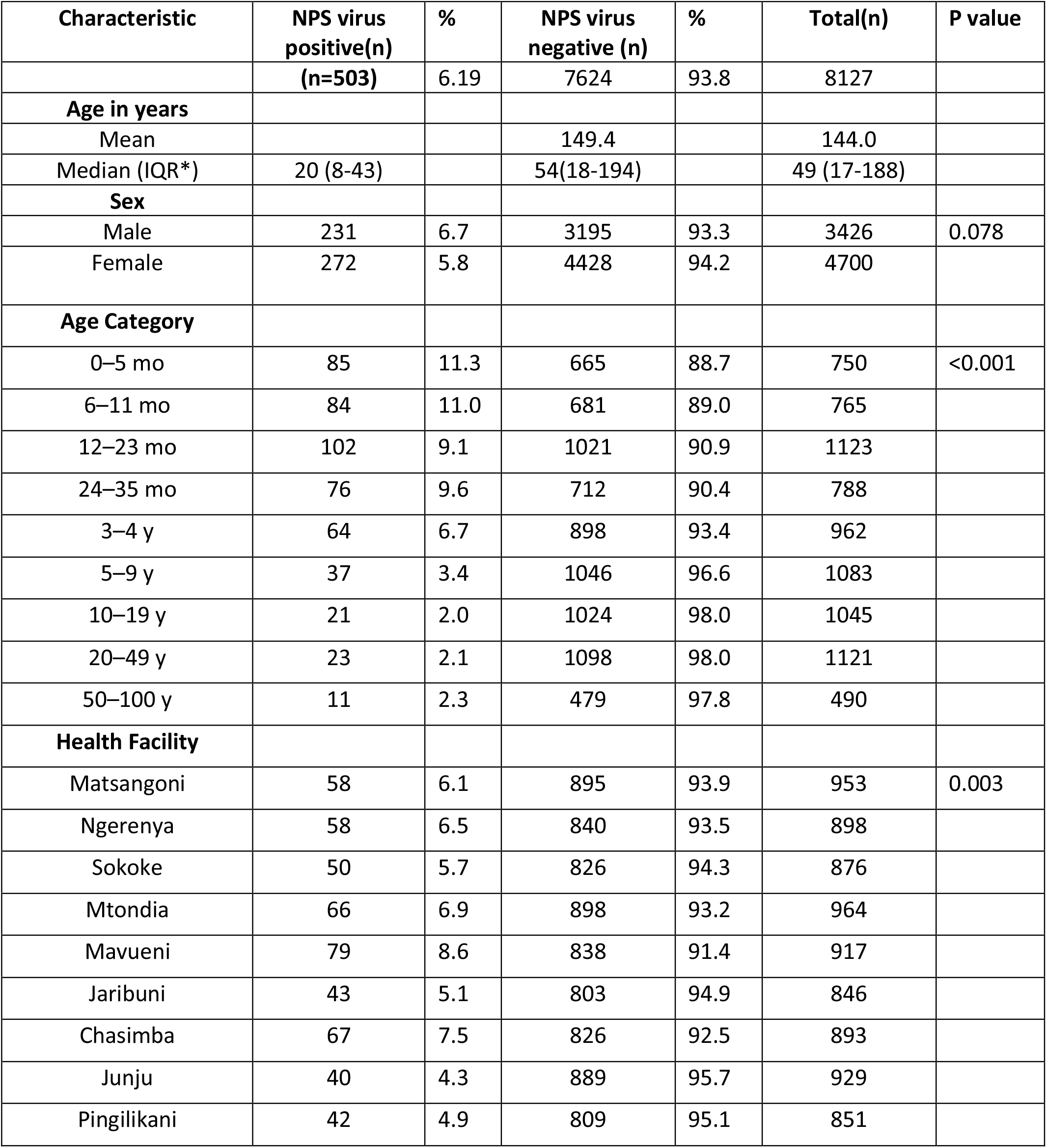
Characteristics of RSV positive and negative study participants by age, gender, and the 9 outpatient health facilities in Kilifi County, coastal Kenya.

| Characteristic | NPS virus positive(n) | % | NPS virus negative (n) | % | Total(n) | P value |
| --- | --- | --- | --- | --- | --- | --- |
|  | (n=503) | 6.19 | 7624 | 93.8 | 8127 |  |
| **Age in years** |  |  |  |  |  |  |
| Mean |  |  | 149.4 |  | 144.0 |  |
| Median (IQR*) | 20 (8-43) |  | 54(18-194) |  | 49 (17-188) |  |
| **Sex** |  |  |  |  |  |  |
| Male | 231 | 6.7 | 3195 | 93.3 | 3426 | 0.078 |
| Female | 272 | 5.8 | 4428 | 94.2 | 4700 |  |
| **Age Category** |  |  |  |  |  |  |
| 0–5 mo | 85 | 11.3 | 665 | 88.7 | 750 | <0.001 |
| 6–11 mo | 84 | 11.0 | 681 | 89.0 | 765 |  |
| 12–23 mo | 102 | 9.1 | 1021 | 90.9 | 1123 |  |
| 24–35 mo | 76 | 9.6 | 712 | 90.4 | 788 |  |
| 3–4 y | 64 | 6.7 | 898 | 93.4 | 962 |  |
| 5–9 y | 37 | 3.4 | 1046 | 96.6 | 1083 |  |
| 10–19 y | 21 | 2.0 | 1024 | 98.0 | 1045 |  |
| 20–49 y | 23 | 2.1 | 1098 | 98.0 | 1121 |  |
| 50–100 y | 11 | 2.3 | 479 | 97.8 | 490 |  |
| **Health Facility** |  |  |  |  |  |  |
| Matsangoni | 58 | 6.1 | 895 | 93.9 | 953 | 0.003 |
| Ngerenya | 58 | 6.5 | 840 | 93.5 | 898 |  |
| Sokoke | 50 | 5.7 | 826 | 94.3 | 876 |  |
| Mtondia | 66 | 6.9 | 898 | 93.2 | 964 |  |
| Mavueni | 79 | 8.6 | 838 | 91.4 | 917 |  |
| Jaribuni | 43 | 5.1 | 803 | 94.9 | 846 |  |
| Chasimba | 67 | 7.5 | 826 | 92.5 | 893 |  |
| Junju | 40 | 4.3 | 889 | 95.7 | 929 |  |
| Pingilikani | 42 | 4.9 | 809 | 95.1 | 851 |  |

**Figure 2.**
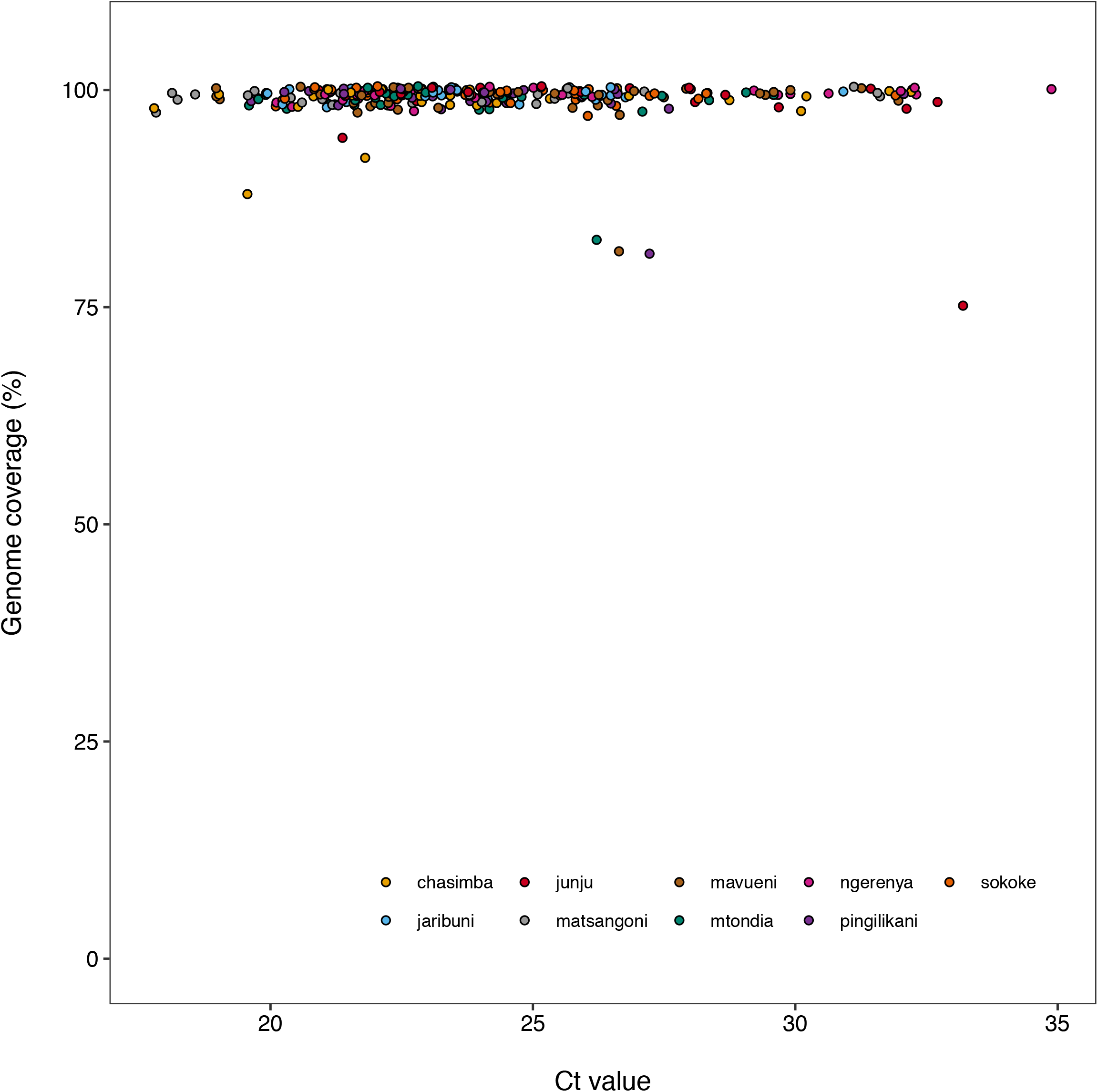
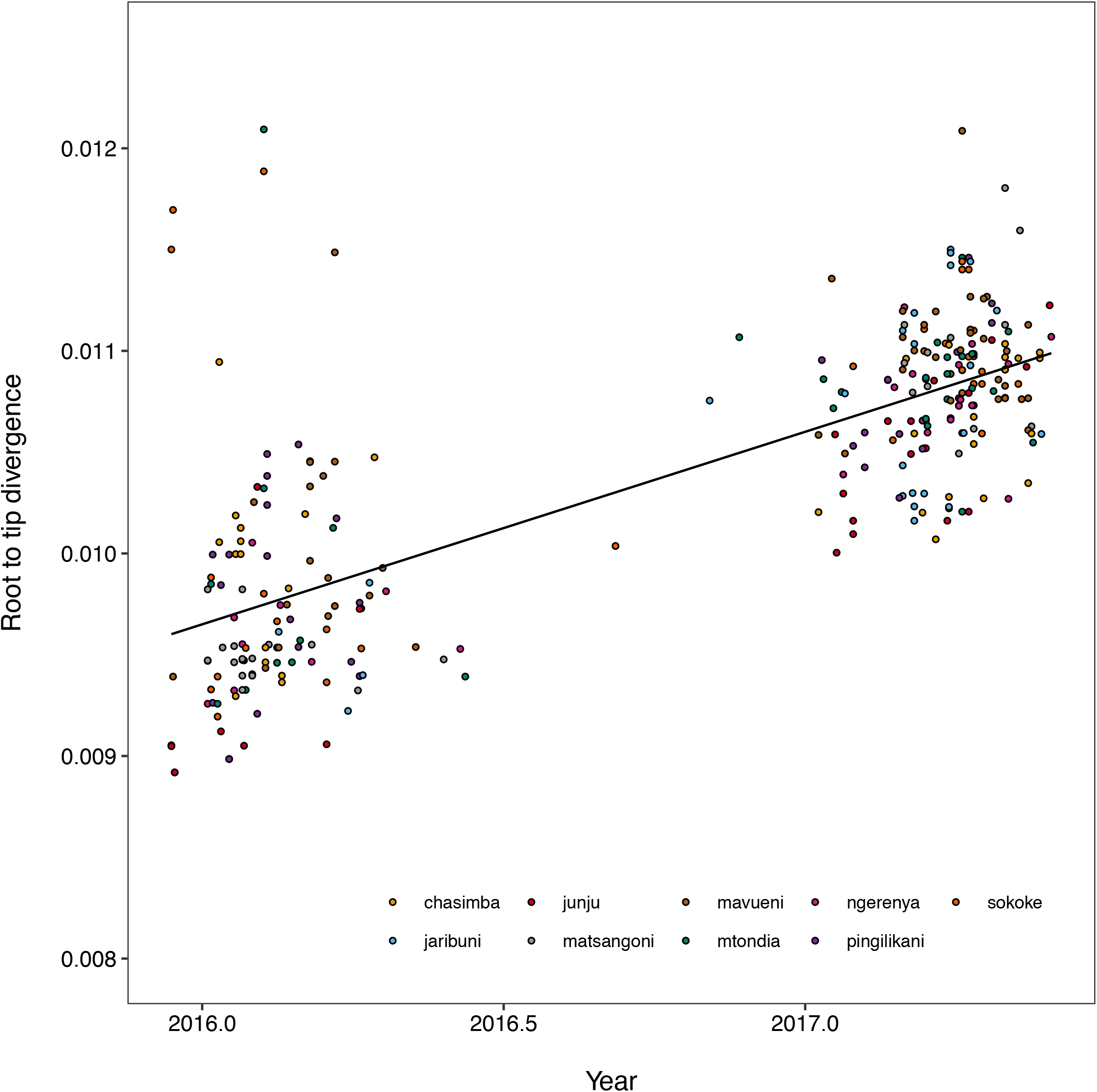
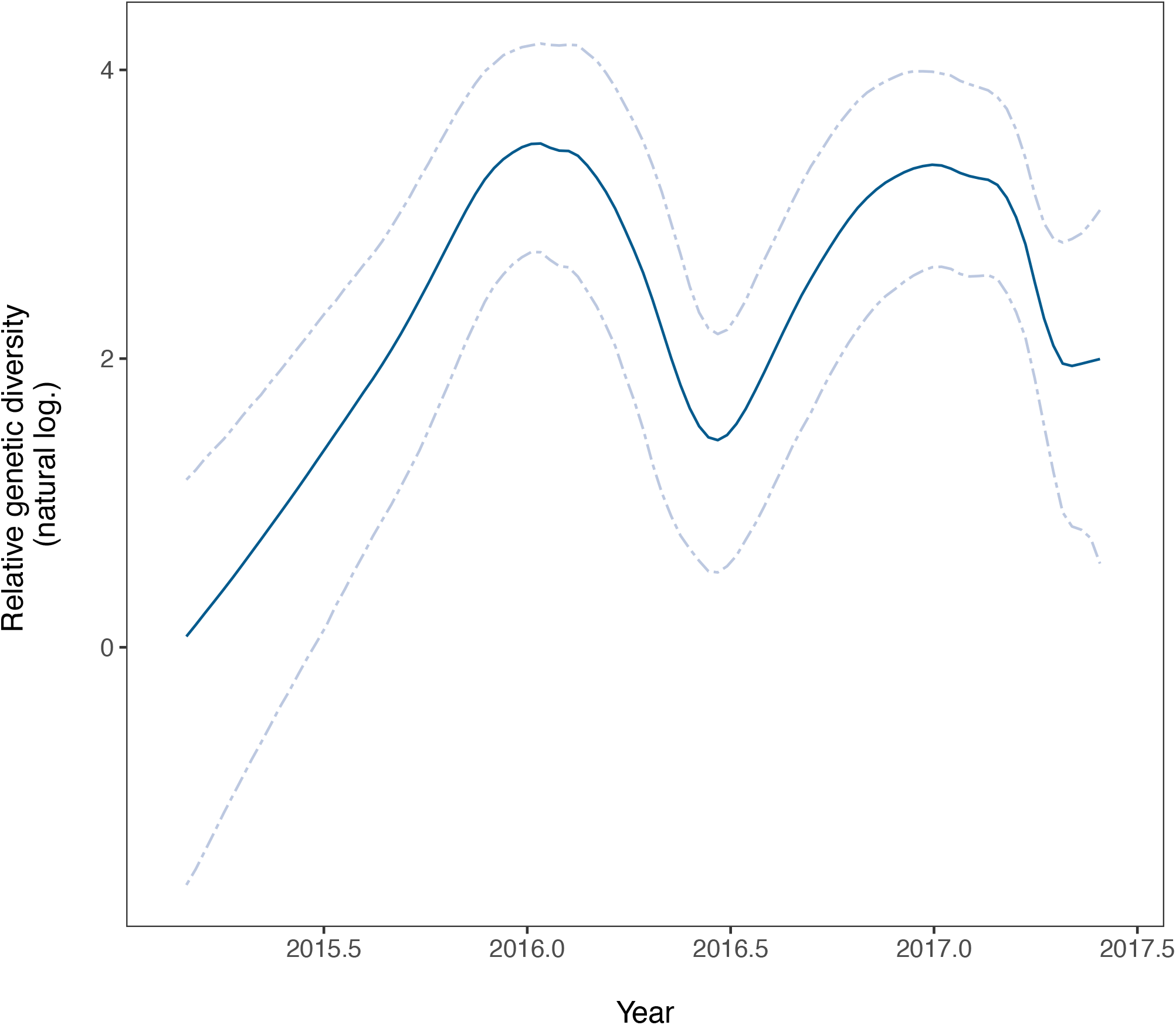
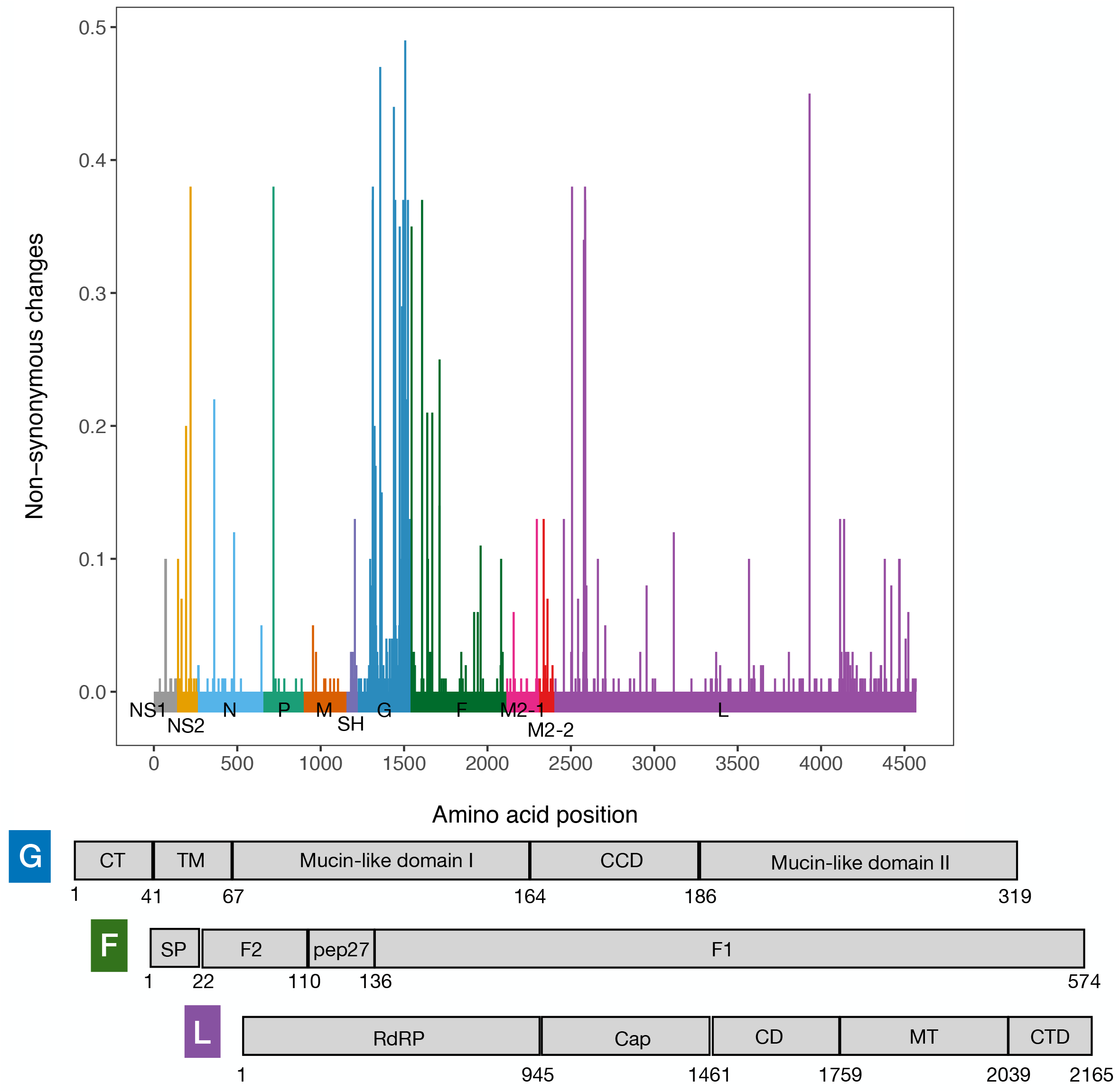
(**A**) Genome coverage for each virus isolate versus the viral load (rRT-PCR Cycle threshold (Ct) value). (**B**) Root-to-tip divergence as a function of sampling time colored by study location. (**C**) Relative genetic diversity through time estimated using the Gaussian Markov Random Field (GMRF) Skygrid model. Solid line represents mean relative genetic diversity while the corresponding dashed lines indicate the 95% HPD intervals. (**D**) Relative frequencies of potential non-synonymous changes across codon-aligned RSV genome sequences. The frequencies for each codon position are calculated as the number of non-synonymous nucleotide substitutions for all pairwise comparisons in a sequence alignment, while excluding ambiguous bases or insertions. Abbreviations: CT = cytoplasmic, TM = transmembrane, CCD = central conserved domain; SP = signal peptide; RdRp = RNA dependent RNA polymerase, Cap = capping, and MT = methyltransferase, CD = connector domain, CTD = C-terminal domain.

RSV co-infection with other respiratory viruses was observed in 41/503 (8.2%) samples: rhinovirus (n =18) and adenovirus (n=11) were co-detected most frequently with RSV. Comparative RSV incidence data was obtained from a contemporaneous respiratory virus surveillance study that recruits patients aged <59 months admitted with acute lower respiratory tract infection at the pediatric ward of the Kilifi County Hospital (KCH, see Supplementary Methods). The frequency of RSV occurrence among children <5 years of age differed significantly (p-value <0.001) between the outpatient and inpatient setting during the period of December 2015 to June 2017. The RSV viral load was similar between the inpatient and outpatient settings (**Supplementary Figure 1(B)**). Across both inpatient and outpatient care settings, the peak time for RSV case detections occurred from November to May the following year (**Supplementary Figure 3**). The distribution of clinical symptoms among patients attending the nine outpatient health facilities, and by age and gender are shown in **Supplementary Table 2**.

**Figure 3.**
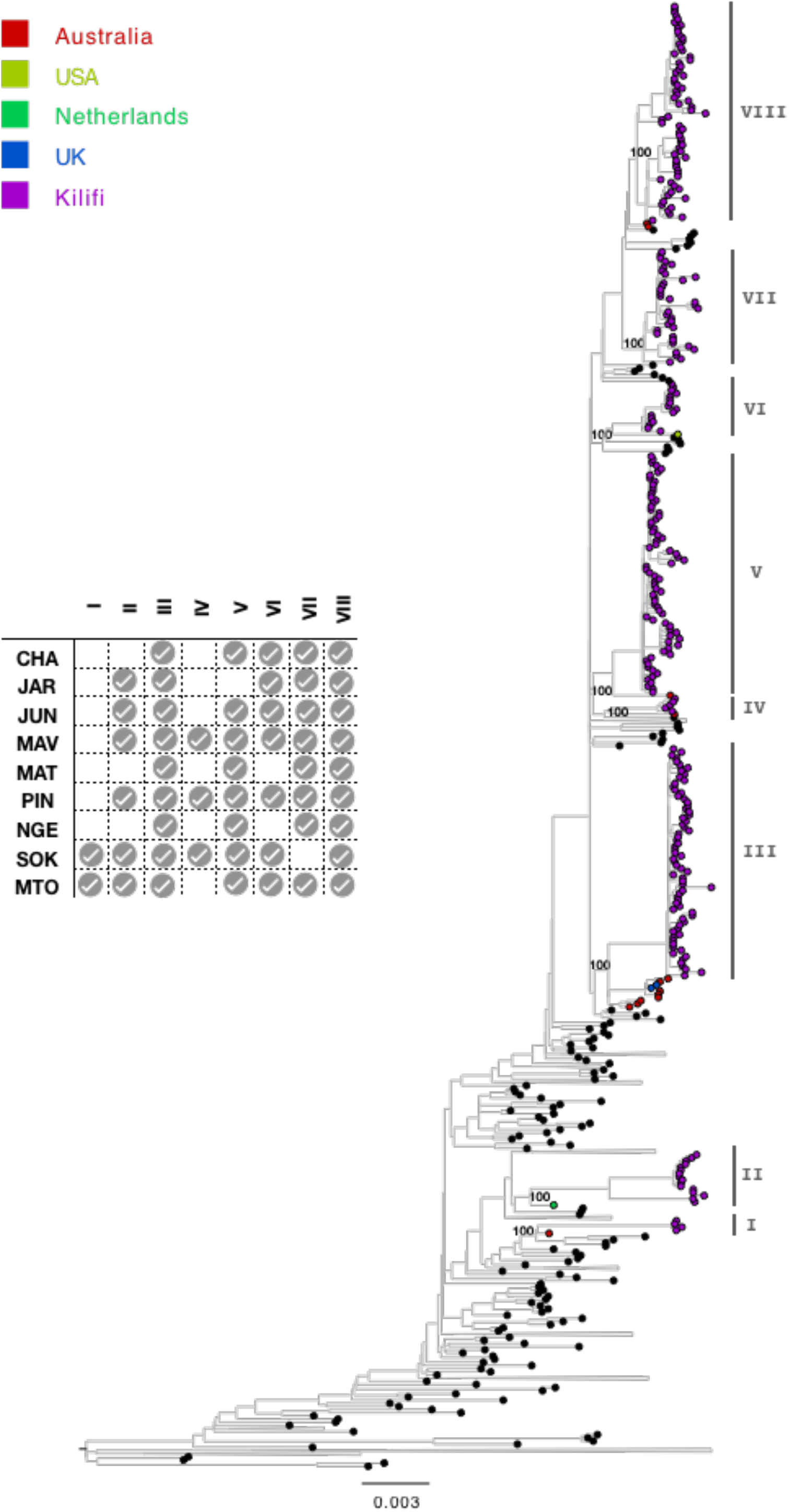
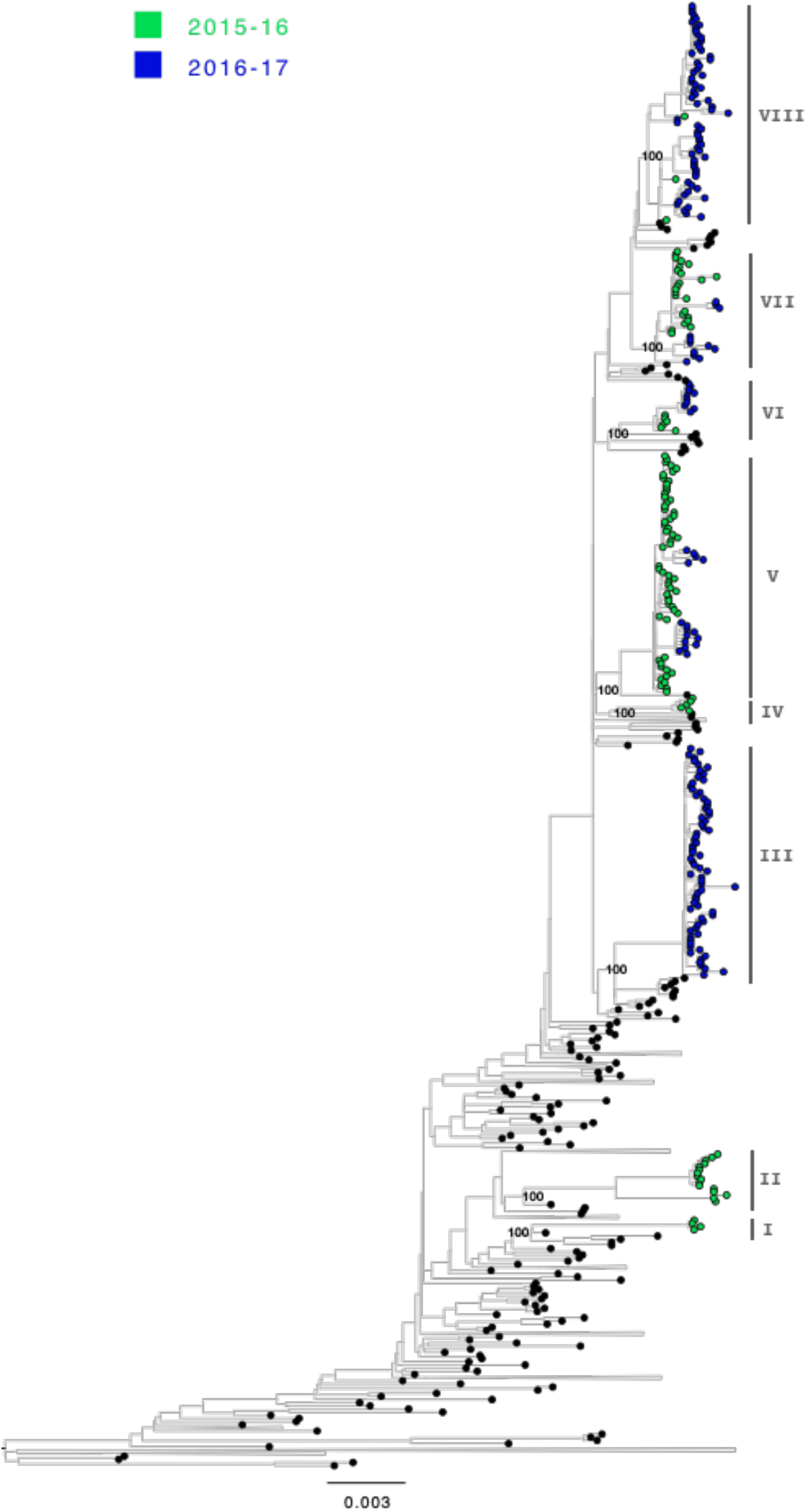
(**A**) Phylogenetic placement of RSVB genomes from Kilifi relative to sequences from other geographical regions. The clade or introduction assignments are indicated as I to VIII on the trees and the circles at the branch tips are colored by location or country of origin. Only the contemporaneous (global) sequences that are closely related to or share common ancestors with RSVB genomes from Kilifi, are colored. The black circles represent other global sequences used in the phylogenetic analysis. Geographic distribution of the RSVB introductions in Kilifi is shown in the inset table. Abbreviations: CHA = Chasimba, JAR = Jaribuni, JUN = Junju, MAV = Mavueni, MAT = Matsangoni, PIN = Pingilikani, NGE = Ngerenya, SOK = Sokoke, MTO = Mtondia. (**B**) Maximum likelihood phylogeny (1000 bootstrap resampling) showing the temporal distribution of the RSVB introductions in Kilifi. The individual sequences from Kilifi are colored by respective RSV epidemic. The black circles represent other global sequences used in the phylogenetic analysis. Bootstrap support values are shown for the most basal nodes of the inferred introductions.

### RSVB genome sequences from Kilifi

WGS and data assembly was successful for 299/408 (73.3%) RSVB positive samples collected between December 2015 and June 2017 from the 9 selected health facilities in Kilifi county, Kenya. The remaining 26% (109/408) sampling fraction were either of low viral load and we could not obtain more than 4 PCR amplicons (81.6%) or were sequenced at insufficient depth or quality for genome assembly (18.4%). 295/299 genomes were coding-complete (all the 11 RSV coding genes were assembled) and were used in subsequent analysis. The median genome length was 15205 (range 11519 to 15257 nt). All the sequenced viruses belonged to the RSVB BA genotype, characterized by the presence of 60-nt duplication in the C-terminal region of the G glycoprotein gene. Genome coverage did not vary by rRT-PCR Ct value (**Figure 2(A)**).

### Genome-wide sequence diversity and evolution

The sequence data showed linear relationship between genetic change and time (root-to-tip correlation coefficient of 0.82, **Figure 2(B)**); a temporal signal that supported the use of molecular clock models. Bayesian analyses estimated an evolutionary rate of 1.063 ⨯ 10^−3^ (95% HPD: 9.2422 ⨯ 10^−4^ – 1.2063 ⨯ 10^−3^) nucleotide substitutions/site/year, and the genetic diversity traced back to a common ancestor dated in 2014 (date in decimal format: 2014.89; 95% HPD: 2014.63 – 2015.14). Demographic reconstruction (**Figure 2(C)**) showed seasonal periodicity in relative genetic diversity that broadly mirrored RSVB incidence in the two epidemics. The peak genetic diversity occurred in January followed by an inter-epidemic bottleneck that indicated lineage or variant displacement events between epidemics.

The Kilifi genome sequences contained 838 consensus level non-gap single nucleotide polymorphisms (SNPs), 554 (66%) of which were parsimony informative. 503/554 (91%) SNPs were located within coding regions, of which 332/503 (66%) were non-synonymous and non-uniformly distributed across the RSV genome. Non-synonymous changes peaked in density at the mucin-like domains of G gene; in the N-terminal of fusion (F) gene; as well as in the N-and C-terminals of RNA-dependent RNA polymerase (L) gene (**Figure 2(D)**).

Selection pressure analyses showed that G and SH glycoproteins had higher global non-synonymous (dN)/synonymous (dS) substitution rate ratio estimates than all other genes (**Table 2**). SLAC analyses identified three amino-acid sites (135, 217 and 285) in G gene under significant positive selection (*P* <0.1). In addition, we used MEME and FUBAR methods to identify codons under pervasive and episodic (diversifying) positive selection. MEME analyses detected 3 diversifying codons in the F gene, and 11 in the G gene (*P* < 0.1) (**Table 2**). The FUBAR method identified 2 codon sites in F gene and 7 in G gene, under episodic positive selection with significant support (posterior probability >0.9) (**Table 2**).

**Table 2.** The predicted nature of selection pressures acting on each genomic region: 1^st^ column shows the computed mean dN/dS rate ratio using SLAC and the 2^nd^ column shows amino-acid sites in F and G gene under episodic selection as identified by MEME analyses. Sites also detected using the FUBAR method, in addition to MEME, are underlined.

| Non-synonymous (dN)/synonymous (dS) substitution rate ratio per site |  | Sites subject to episodic positive/diversifying selection |
| --- | --- | --- |
| **NS1** | 0.12 | *G gene* |
| **NS2** | 0.236 | <u>135</u>, 144, <u>154</u>, 172, 208, <u>217</u>, |
| **N** | 0.0832 | <u>285</u>, 291, <u>294</u>, 298, <u>303</u> |
| **M** | 0.0525 | <u>252</u>* |
| **P** | 0.0642 |  |
| **F** | 0.179 | *F gene* |
| **G** | 0.487 | <u>125</u>, <u>172</u>, 173 |
| **SH** | 0.426 |  |
| **M2-1** | 0.264 |  |
| **M2-2** | 0.267 |  |
| **L** | 0.122 |  |

\* This site was detected by the FUBAR method only.

### Viral introductions and spread within Kilifi

Genomic phylogenetic and phylogeographic analyses afforded an in-depth look into the introduction and spread of RSVB in the study population between December 2015 and June 2017. To determine viral introductions, we analyzed the Kilifi RSVB genomes (*n=295*) in the context of other global RSVB genomes (*n=500*). Sequences from Kilifi fell into eight major clades (numbered I to VIII, **Figure 3(A)**) in which they clustered more closely among themselves and less within the diversity of contemporaneous strains, indicating at least eight separate introductions into the study population. The absence of external/global strains within the eight clades might suggest local diversification of viruses, although we cannot exclude additional importation from unsampled locations. Still, the low availability of RSVB sequence data from other regions within Kenya or neighboring countries makes it difficult to estimate the precise number of viral introductions.

The first four introductions (I-IV) comprised viruses confined to a single RSV epidemic (**Figure 3(B)**). Clades I, II and IV comprised 25 viruses in total, from the 2015/16 epidemic, while clade III comprised 73 viruses uniquely from the 2016/17 epidemic. The other four clades (V-VIII) comprised 76, 16, 36 and 69 viruses, respectively, a mix from both RSV epidemics. All the eight introductions circulated within more than one location (table inset, **Figure 3(A)**).Viruses in clade III in Figure 3(A) were closely related to RSVB strains collected in January and August 2016 in UK and Australia, respectively. Clades I, IV, V and VIII, shared common ancestors with sequences from Australia. Viruses in Clade VI were closely related to a sequence collected in USA in 2015, while those in clade II were closely related to a Dutch strain. There is a strong potential for bias in these results due to heterogeneous sampling globally. We used TreeTime (42) for maximum likelihood dating of the inferred clades, and the most likely dates of introduction were placed between July 2007 and September 2014 (**Supplementary Table 3**).

Analyzed separately, the time measured MCC tree of Kilifi genomes showed phylogenetic clustering by epidemic, which depicted a temporally structured viral population (**Figure 3(B)**). Our analysis of the geographical signal revealed high AI and PS values (**Table 3**), suggesting relatively extensive viral migration dynamics among study locations. The association between phylogenetic clustering and study location was significant (*p* value < 0.001) in at least 8/9 study locations as revealed by the maximum clade size values (**Table 3**). Differences in the observed and expected MC values (**Table 3**) suggested that Mavueni exhibited the most spatial structure (difference of 8.7) and Mtondia had the least (difference of 0.3).

**Table 3.** Results of Bayesian analysis of phylogeographic structure of RSVB viruses in Kilifi, coastal Kenya, 2015-2017. *P* values correspond to the proportion of trees from the expected (null) distribution equal to, or more extreme than, the median posterior of the statistic. Abbreviations: CHA = Chasimba, JAR = Jaribuni, JUN = Junju, MAV = Mavueni, MAT = Matsangoni, PIN = Pingilikani, NGE = Ngerenya, SOK = Sokoke, MTO = Mtondia.

| Location | Association Index (AI) |  |  | Parsimony Score (PS) |  |  | Maximum Clade size |  |  |  |
| --- | --- | --- | --- | --- | --- | --- | --- | --- | --- | --- |
|  | (95% CI) † |  |  | (95% CI) † |  |  | (95% CI) ‡ |  |  |  |
|  | Observed | Expected | *P* value | Observed | Expected | *P* value | Observed | Expected | *P* value | Difference § |
| ALL | 14.8 (13.8-15.8) | 31 (29.5-32.3) | 0.0 | 132.6 (129-136) | 208.4 (202-214.2) | 0.0 | - | - | - | - |
| CHA | - | - | - | - | - | - | 4.6 (4-6) | 1.84 (1.1-2.7) | 10E-4 | 2.62 |
| JAR | - | - | - | - | - | - | 5.27 (5-6) | 1.56 (1-2.08) | 10E-4 | 3.71 |
| JUN | - | - | - | - | - | - | 6 (6-6) | 1.55 (1-2.03) | 10E-4 | 4.45 |
| MAT | - | - | - | - | - | - | 6.32 (4-10) | 1.71 (1-2.2) | 10E-4 | 4.61 |
| MAV | - | - | - | - | - | - | 11 (11-11) | 2.3 (1.76-3) | 10E-4 | 8.7 |
| MTO | - | - | - | - | - | - | 2 (2-2) | 1.7 (1-2.3) | 0.21 | 0.3 |
| NGE | - | - | - | - | - | - | 3.65 (2-4) | 1.64 (1-2.2) | 10E-4 | 2.01 |
| PIN | - | - | - | - | - | - | 3.1 (3-4) | 1.73 (1-2.2) | 0.0084 | 1.37 |
| SOK | - | - | - | - | - | - | 4 (3-5) | 1.8 (1.1-2.3) | 10E-4 | 2.2 |

† AI and PS metrics were determined for all locations combined.
‡ Maximum clade size was determined for each specific location.
§ Difference between observed and expected (null) clade size.

We compared the model fit for symmetrical (reversible) and asymmetrical (non-reversible) discrete diffusion models with BSSVS procedure. The asymmetrical CTMC model gave a better fit (marginal likelihood, path sampling = −264.14 and stepping-stone sampling = −316.3) compared to the reversible’s marginal likelihood path sampling of −287.44 and stepping-stone sampling of −351.9. The non-reversible model is a more realistic description of the diffusion process that allows location exchange rates to vary according to directionality (37). Mavueni may have played a central role in virus dissemination to other study locations: it was the most probable location for most ancestral nodes with location posterior support >0.9, (**Figure 4(A)**), and viral lineage movements from Mavueni were statistically supported with Bayes Factor >1000 (**Table 4**). In addition, there was a relatively high genomic diversity in Mavueni (red-colored taxa in **Supplementary Figure 4**). The inferred viral movements between the nine study locations are visualized as geographic links in **Figure 4(B)**. There were very few statistically supported viral movements between Sokoke and other study locations.

**Table 4.** Statistically supported state transitions indicating viral migration events. Bayes factor >6 was considered significant.

| Transition between |  | Distance (Km)<sup>Φ</sup> | Bayes factor | Mean indicator <sup>δ</sup> | Mean rate |
| --- | --- | --- | --- | --- | --- |
| Chasimba | Mtondia | 21.5 | 6.2 | 0.46 | 1 |
| Jaribuni | Junju | 17.5 | 6.7 | 0.48 | 1 |
| Chasimba | Ngerenya | 25.5 | 8.3 | 0.53 | 0.8 |
| Soko | Matsangoni | 20.6 | 9.8 | 0.57 | 0.9 |
| Mtondia | Junju | 24 | 12.2 | 0.63 | 0.76 |
| Chasimba | Pingilikani | 6.2 | 13.7 | 0.65 | 0.9 |
| Pingilikani | Mtondia | 21.6 | 18.2 | 0.71 | 1 |
| Junju | Jaribuni | 17.5 | 20.8 | 0.74 | 1 |
| Mavueni | Mtondia | 12.5 | 28.5 | 0.80 | 1 |
| Matsangoni | Ngerenya | 12.3 | 35.0 | 0.83 | 0.75 |
| Mavueni | Matsangoni | 29.3 | 179.9 | 0.96 | 0.87 |
| Mavueni | Junju | 11.5 | 434.6 | 0.98 | 0.87 |
| Junju | Pingilikani | 5.4 | 1166.6 | 0.99 | 1.4 |
| Ngerenya | Mtondia | 4.6 | 1624.5 | 1.00 | 1.3 |
| Mavueni | Ngerenya | 17 | 2062.5 | 1.00 | 1.2 |
| Soko | Mtondia | 10.9 | 12678.4 | 1.00 | 1.7 |
| Mavueni | Jaribuni | 12.8 | 24571.2 | 1.00 | 1.09 |
| Mavueni | Pingilikani | 9.8 | 30243.2 | 1.00 | 1.5 |
| Mavueni | Chasimba | 12.1 | 196620.8 | 1.00 | 1.75 |
| Mavueni | Soko | 16.5 | 393248.8 | 1.00 | 1.78 |

<sup>Φ</sup> Great circle distance estimates between centroid longitude and latitude of the respective locations.
<sup>δ</sup> The posterior probability of observing non-zero migration rates in the Bayesian sampled trees.

**Figure 4.**
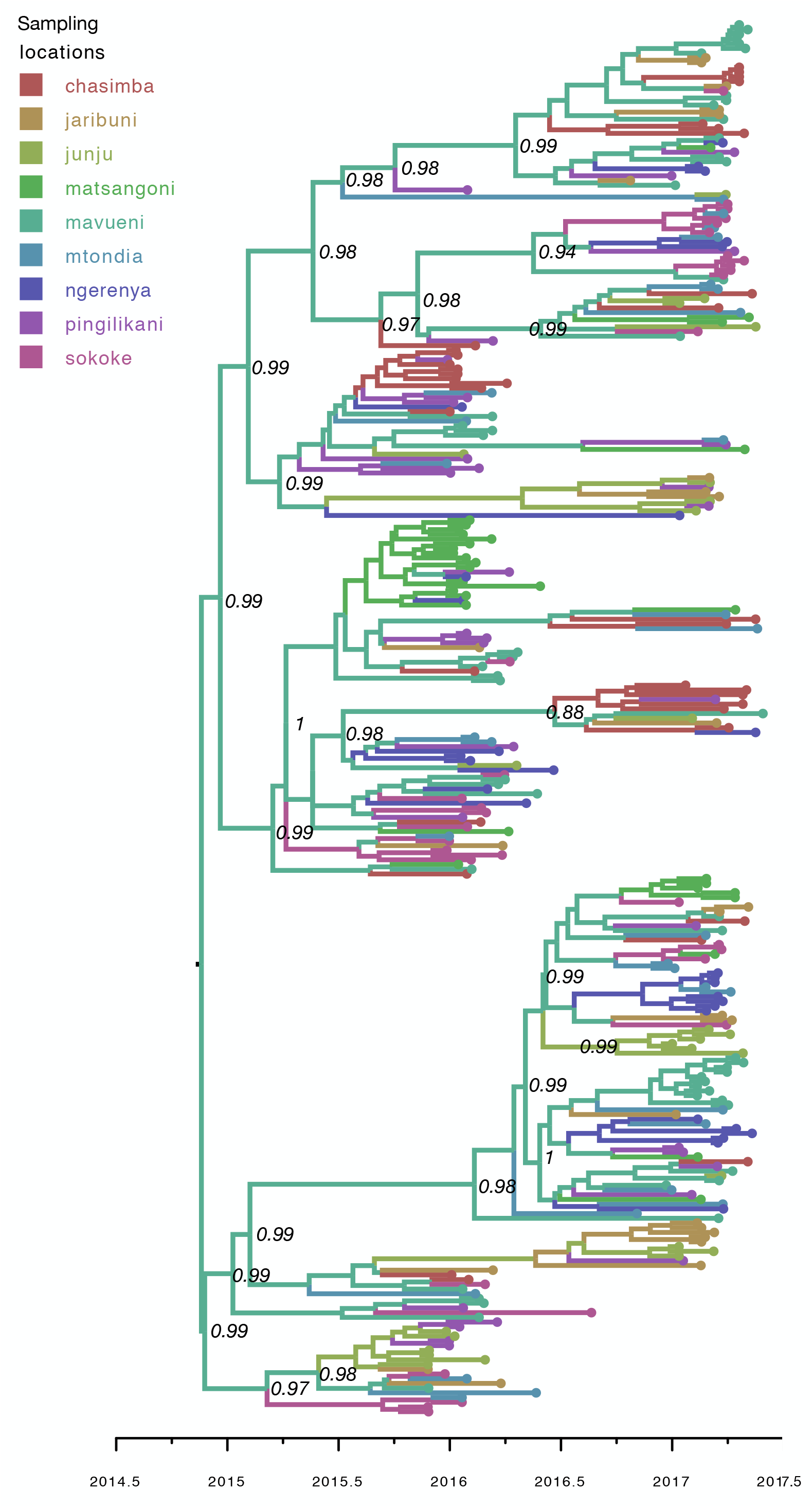
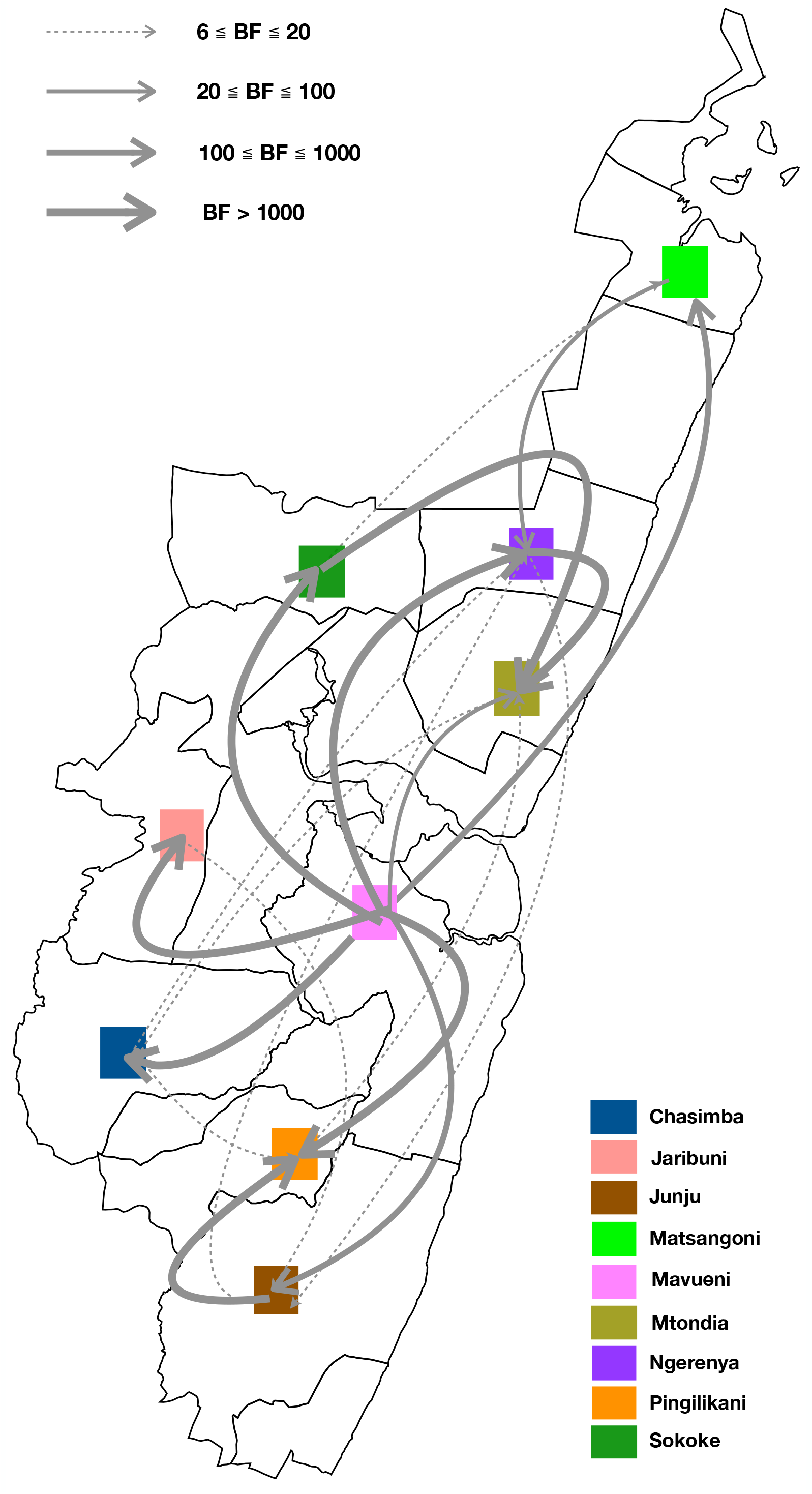
(**A**) Temporal-scaled phylogeographic maximum clade credibility tree of Kilifi RSVB genomes. Branch colors indicate the modal location (most probable reconstructed location at each node) inferred under the asymmetrical discrete phylogeographic model. Location-state posterior probabilities are shown next to relevant nodes along with the posterior support. (**B**) Spatial diffusion pathways of RSVB in Kilifi. Well supported (Bayes Factor >6) discrete diffusion asymmetric rates of viral movement between the study locations are indicated. The type and thickness of the arrows represents the relative strength of the diffusion rate.

### Phylogenomic clusters

The RSVB viruses in Kilifi were further summarized as phylogenomic clusters based on genetic similarity of consensus genomes. For this, we examined the distribution of maximum likelihood pairwise evolutionary distances among the 295 samples (43365 pairs). The genetic distance distribution was multi-modal (**Figure 5 (A)**), and we defined a phylogenomic cluster as a group of viruses (≥2) within an epidemic that contained evolutionary pairwise distances of <0.0024 nucleotide substitutions per site (red dashed line **Figure 5 (A)**) or 37 pairwise nucleotide differences. The distance threshold of <0.0024 nucleotide substitutions per site demarcated the 17^th^ percentile of the entire distance distribution and was additionally supported by clear separation of viruses into closely related phylogenetic clusters with >0.9 posterior probability support.

**Figure 5.**
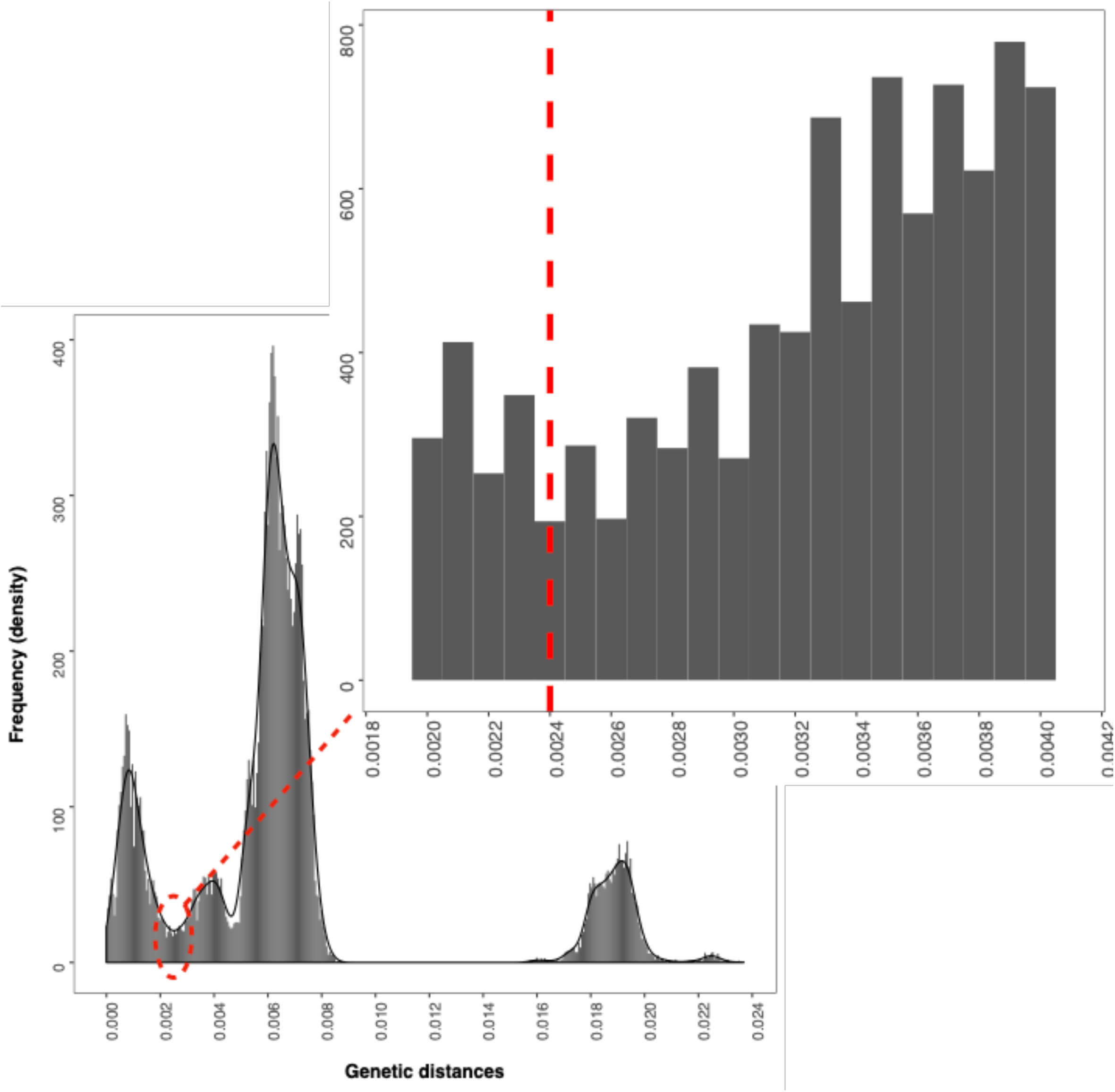
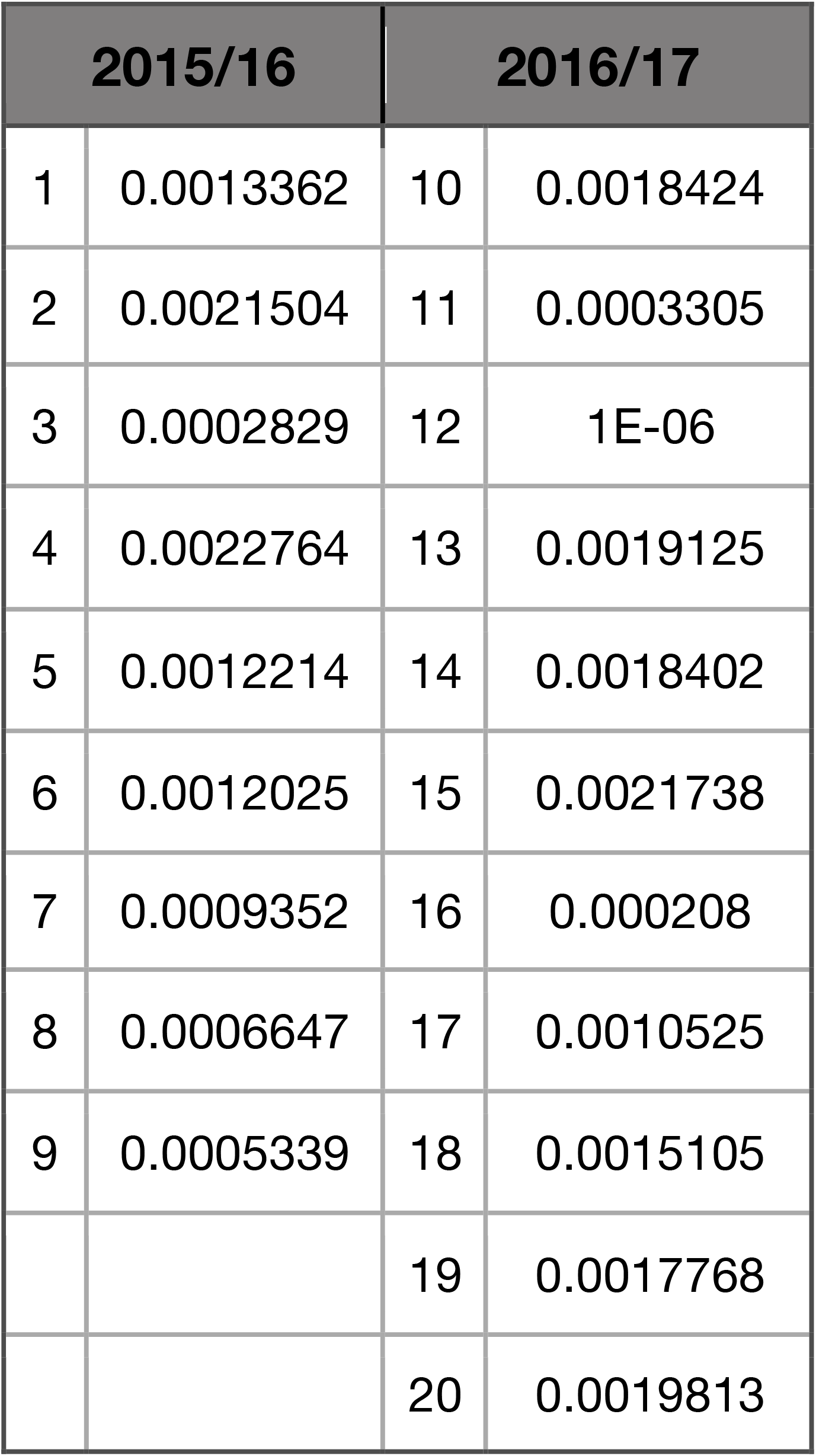
(**A**) Histogram of the whole genome sequences patristic distance frequency distribution. The vertical red dashed line corresponds to the 17^th^ percentile distance threshold (0.0024 expected nucleotide substitutions per site) for which phylogenomic clusters were identified. The distances were measured in units of nucleotide substitutions per site and extracted from a maximum likelihood phylogeny (1000 bootstrap resampling). (**B**) Maximum pairwise patristic distances for sequences belonging to the different phylogenomic clusters.

Overall, at least 20 phylogenomic clusters were identified, all of which were supported by strong posterior probability support (>0.9); nine in the 2015/16 epidemic (**Figure 6(A)**) and eleven (**Figure 6(B)**) in the 2016/17 epidemic. The lowest within-cluster genetic diversity was found in cluster 12 in the 2016/17 epidemic and the highest in cluster four in the 2015/16 epidemic (**Figure 5(B)**). Genetically, cluster five in the 2015/16 epidemic fell basal to cluster 17 in the 2016/17 epidemic and both made the 6^th^ RSVB introduction (clade VI in **Figure 3(A)**), a phylogenetic pattern compatible with in-situ virus evolution and persistence. The clusters ranged in size, from 2 to 72 viruses. Overall, there was abundant viral diversity in Pingilikani in both epidemics (13 clusters), while Matsangoni and Ngerenya had the lesser diversity (six clusters). In particular, only one cluster or viral lineage was circulating in Matsangoni in the 2015/16 epidemic.

**Figure 6.**
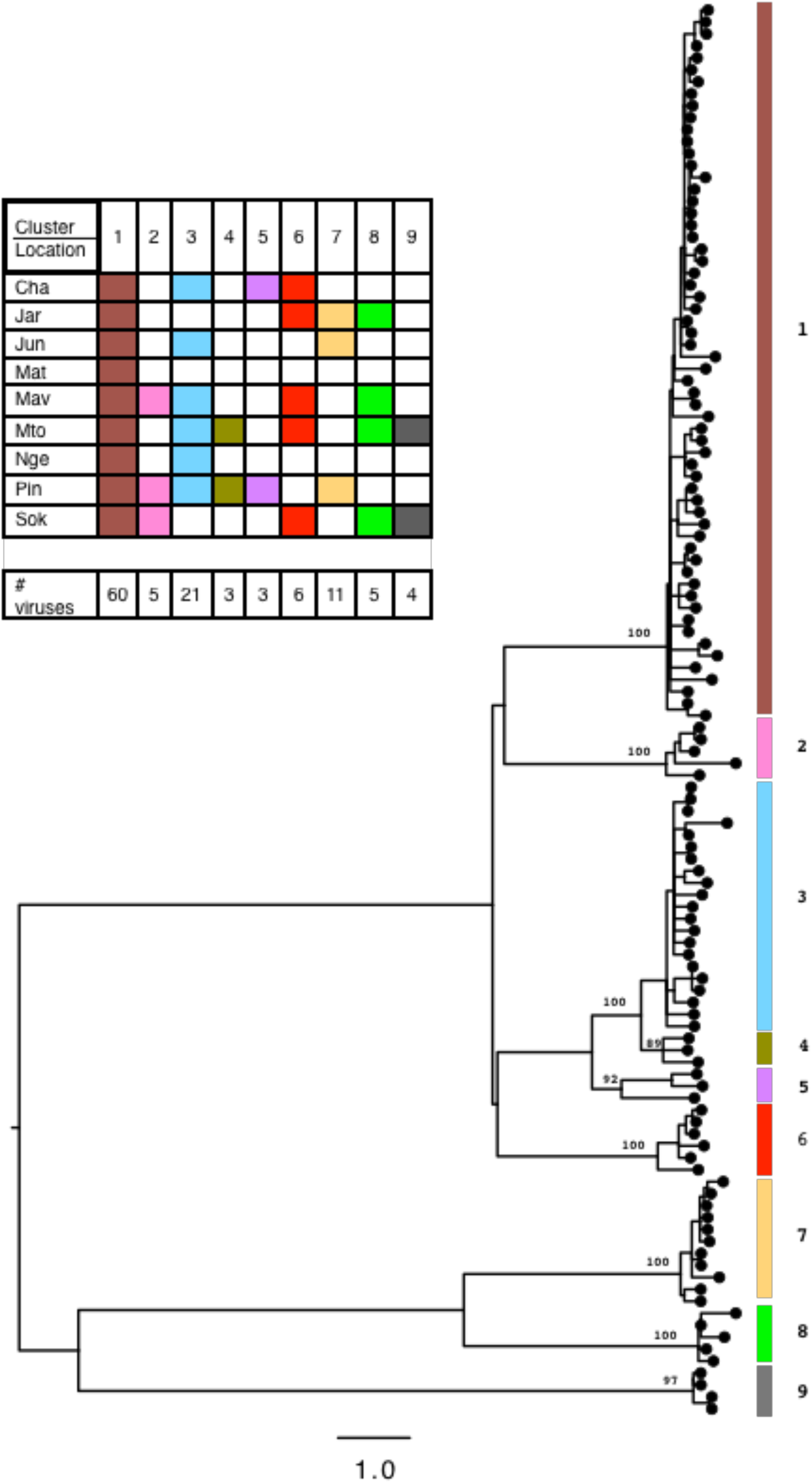
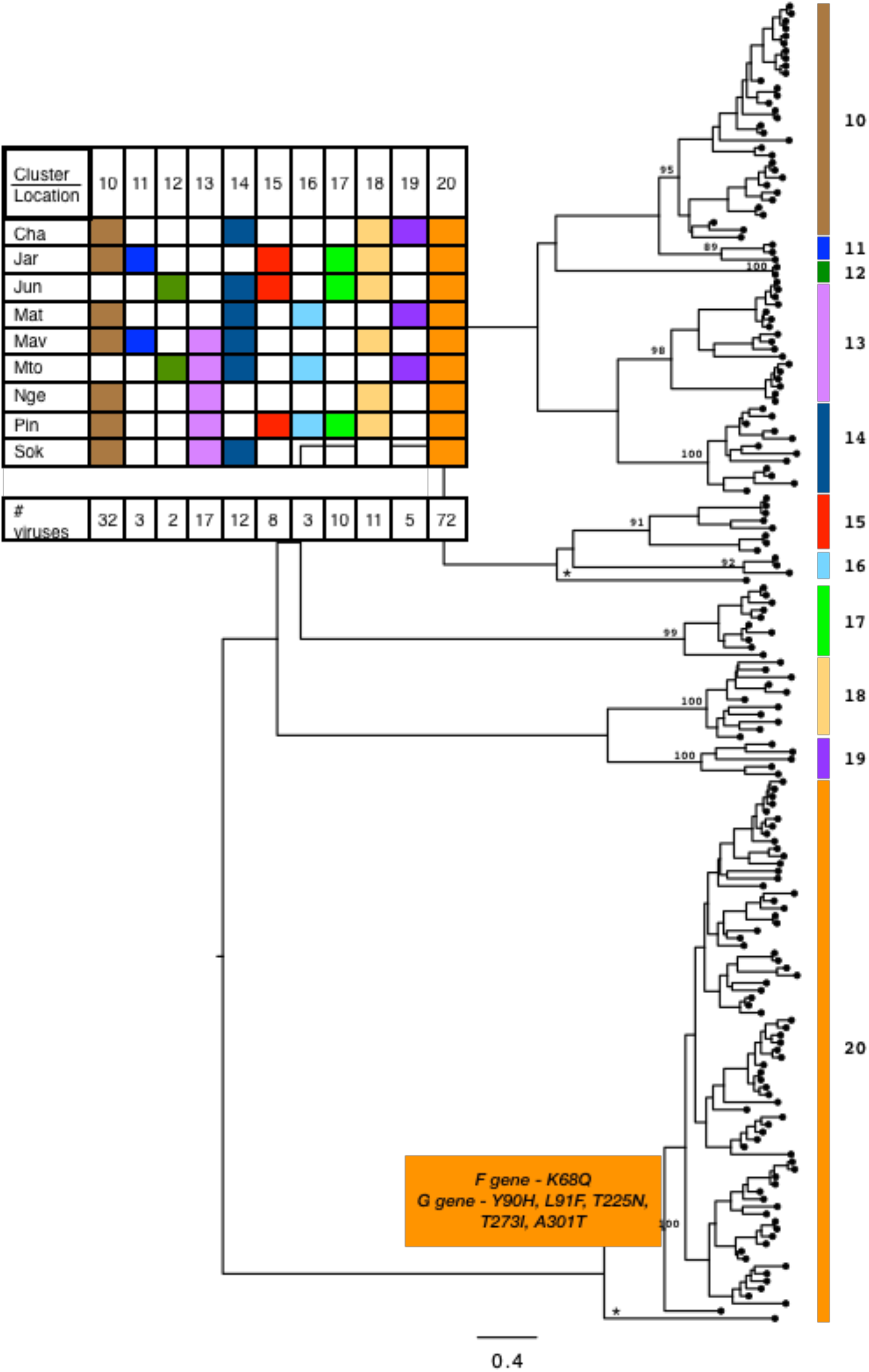
The assigned phylogenomic clusters are shown on maximum likelihood phylogenies (1000 bootstrap resampling): 9 clusters in the 2015/16 epidemic (**A**) and 11 clusters in the 2016/17 epidemic (**B**). Geographical composition and number of viruses/sequences per phylogenomic cluster are shown (inset tables). The ‘*’ in (B) indicates sequences that were not assigned into a phylogenomic cluster. All sequences had epidemiological and geographical information. Amino-acid changes in F and G glycoproteins unique to the emergent variant (cluster 21) in the 2016/17 are labeled on the major branch leading to the cluster. Scale bar represents the number of substitutions per site.

Sample collection dates were used to estimate the duration of each phylogenomic cluster. Clusters 20 (172 days), 10 (173 days) and six (211 days) circulated over a wide temporal scale up to several months. Geographically, clusters one and 20 were the largest in size and most pervasive circulating widely in all the nine study locations (**Figure 6**). Two viruses in the 2016/17 epidemic (marked with ‘*’ in **Figure 6**) were singletons and could not be assigned to any phylogenomic cluster, likely an artifact of limited sampling. The number of clusters identified in each study location was weakly associated with the number of genomes from that location (Spearman correlation *rho* = 0.2, *P* = 0.6)

### Amino acid diversity in F and G genes

Given their role in the initial phases of infection and as the major antigens for eliciting neutralizing antibody responses, we examined amino-acid variation in the two major glycoproteins on the surface of the RSV virion, the attachment (G) glycoprotein and the fusion (F) glycoprotein. The F protein is also the major target for antiviral immuno-prophylaxis. In all the sequenced viruses, there were 37 and 93 amino-acid (codon) sites containing non-synonymous changes in F and G gene, respectively, and the frequency of the observed non-synonymous variants was up to 24.7% and 90.4% in F and G gene, respectively. While most of these variable sites contained a single non-synonymous change, one codon in F contained two changes (K419N/R), and three codons in G contained multiple non-synonymous changes (T107A/D, P289L/S and E290A/G/V). The K419N and K419R substitutions were seen in a subset (n=6) of sequences in cluster 13 and in cluster 17, respectively. The T107D substitution was observed in a subset (n=7) of sequences in cluster 13, while the T107A was seen in all other clusters except for seven, eight and nine. The P289L and P289S substitutions were observed in cluster seven and eight, and six, respectively. The E290A, E290G and E290V substitutions were observed in clusters 13, 6 and 4 and 14, respectively. These amino-acid variants were not limited to a study location and were relative to the earliest dated sequence in the dataset (14 December 2015).

In F gene, viruses in clusters 10 and 11 (**Figure 6(B)**) in the 2016/17 epidemic contained two unique amino-acid changes (N99T and T129I). Additionally, three non-synonymous substitutions (V103A, L172Q and S173L) located in the F protein antigenic site V were unique to clusters seven, eight and nine in the 2015/16 epidemic. In G gene, 90% of the genomes contained T107A non-synonymous substitution, 64% had P217L substitution, and 45% contained a H285Y substitution, compared to the earliest sequence. A number of amino-acid changes occurred at the node between cluster nine and other viruses (marked A in **Figure 3(A)**) and these included N213D, A227I, P255S, and N303E in G gene; and T19A and P125L in F gene. A non-synonymous change from <u>C</u>AA(Q) resulted in early Stop codon (<u>T</u>AA) at amino acid position 311 in 135/295 (45%) sequenced viruses.

Several amino-acid sites appeared to undergo reversal mutation, for instance in G gene, T/S75S/T occurred within the 2015/16 epidemic, while T/I252I/T, L/P270P/L and G/S135S/G occurred in both epidemics. The I/L542L/I reversal mutation in F gene was seen in the 2015/16 epidemic. The T/S75S/T (G gene) and I/L542L/I (F gene) reversal substitutions were both observed in phylogenomic clusters seven and eight that comprised clade II (**Figure 3(A)**). The L/P270P/L reversal mutation occurred in clusters 5, 7 and 17. Reversible evolution may contribute to the escape from the human population immune response, thereby facilitating viral transmission (43).

### Emerging genetically distinct variant in 2016/17 epidemic

Several non-synonymous changes were unique to phylogenomic cluster 20 in the 2016/17 epidemic (**Figure 6(B)**), including K68Q in F gene; Y90H, L91F, T225N, T273I, and A301T in G gene. These six non-synonymous changes were co-occurring in all the sequences in cluster 20 and were detected at an intermediate frequency (41% of samples in 2016/17) in the study population.

Importantly, the amino-acid change K68Q is at the antigenic site Ø, the binding site of monoclonal antibody (mAb) MEDI8897. A variant with the mutation K68N was previously detected in 2% of sampled viruses in the US (44). Other amino-acid changes distinctive to the cluster 20 are listed in **Supplementary Table 4**. This phylogenomic cluster, which was also unequivocally an independent introduction (clade III, **Figure 3(A)**), was the most prevalent and its detection through genomic analysis rules out any sample contamination and sequencing errors.

We went further and checked whether this emerging variant was also present in the wider Kilifi county based on G gene sequence data of samples collected from pediatric ward admissions (<59 months of age) at the Kilifi County Hospital (KCH, formerly Kilifi District Hospital) during the 2016/17 RSV epidemic. KCH provides primary care and inpatient referral services to a larger catchment area. We found that 32% of RSVB positive hospital samples in the 2016/17 epidemic clustered with the emerging variant and likewise contained the non-synonymous changes Y90H, L91F, T225N, T273I, and A301T in G gene, which further increased the overall prevalence of clade III in the 2016/17 epidemic.

We also investigated whether the emerging variant persisted in Kilifi after the 2016/17 RSV epidemic, based on G gene sequences of RSVB positive samples collected from pediatric admissions at KCH during the 2018/19 epidemic. To note, there were no RSVB cases in the 2017/18 epidemic in the KHDSS study (outpatient surveillance in the nine health facilities) (**Supplementary Figure 5**) and only 2 RSVB cases were detected in the in-patient pediatric admissions at KCH (<0.01% of RSV positive samples). We found that viruses in 2018/19 epidemic clustered closely with clade III or cluster 20 and the branch leading to the 2018/19 RSVB epidemic shared a common ancestor with cluster 20 (**Supplementary Figure 6**). The 2018/19 viruses likely descended from the same introduction as clade III, suggesting that the mutations defining clade III were epidemiologically important. It is possible that RSVB underwent a significant population (or genetic) bottleneck after the 2016/17 epidemic and older viruses were eliminated given that no RSVB viruses or persistent variants were transmissible in the 2017/18 epidemic.

### Amino-acid diversity in other regions of the genome

Additional variable amino acid sites observed in the other genomic regions are listed in (**Supplementary Table 4**).

Prominent amino acid changes occurring in more than one phylogenomic cluster are underlined in **Supplementary Table 4**. The emerging variant (cluster 20) discussed above had two other unique amino acid differences in NS2 (non-structural protein 2) and L (polymerase) genes. Even in other genomic regions, phylogenomic clusters 5 (2015/16 epidemic) and 17 (2016/17 epidemic), which are nested within the same virus introduction (clade VI, **Figure 3(A)**), shared amino-acid differences. Clusters 10 to 14 had the same amino-acid differences in F and L genes.

## Discussion

Here we report whole genome molecular epidemiology and phylodynamics of respiratory syncytial virus group B providing a detailed view of the introduction and spread the virus in Kilifi county, coastal Kenya. The results were obtained from genomic analyses of 295 samples originating from representative sampling across the KHDSS area, over two consecutive RSV epidemics. Phylogenetic analyses revealed multiple virus introductions, each introduction commonly circulating in all the study locations, suggesting substantial spatial spread and transmission between locations in a relatively short time. Although RSV surveillance has improved in many regions across the world, publicly available RSVB genomic data from recent years is quite insufficient and may have limited our inference of spatial and temporal placement of virus introductions in Kilifi.

Analysis of RSVB transmission dynamics in Kilifi suggested extensive viral migrations among the study locations associated with the nine health facilities as well as strong spatial substructures within each location. The substructures might represent predominant local transmission and diversification processes. Using the BSSVS approach and an asymmetrical diffusion model to reconstruct RSVB dispersal, we observed significant and strong support for epidemiological links between one central location (Mavueni), located at the intersection of the main roads through the KHDSS and other study locations (BF > 1000, Table 4). However, we note that Mavueni had a larger proportion of RSVB genomes compared to other locations, which might bias the ancestral location estimates. Improved road infrastructure and transportation within KHDSS has facilitated mobility thus increasingly connecting the local population and expanding virus transmission networks.

Temporal sequence divergence and accumulation of nucleotide substitutions was detectable over the sampling timeframe and varied by epidemic intervals as shown by two separate groups of tips in the linear regression plot (Figure 2B). Time-scaled phylogenies also exhibited a marked epidemic behavior indicative of chronological generation of new variants. The two epidemics were characterized by multiple clades for viruses sampled within the same epidemic, indicating continued transmission generated and sustained by increasing spatial connectivity in the wider Kilifi county. Changes in the relative genetic diversity coincided with RSVB case detection and captured fine temporal resolution of changes in the viral population size, which also implied sufficient sampling density (14). While estimating nucleotide substitution rate is useful in revealing the dynamics and processes of viral evolution (45), we could not directly compare our estimate of the mean nucleotide substitution rate with previous studies due to varying sampling timescales, different molecular clock (fixed vs. relaxed) and coalescent models, as well as epidemiologic variations.

Phylogenetic clusters have been used to investigate epidemiologically significant HIV hotspots and characterize groups burdened by a high rate of HIV transmission (46). We inferred 20 phylogenomic clusters of closely related viruses based on genetic relatedness, which we put forward as potential transmission units. Stochastic difference in transmissibility or circulation, or infection rates could explain the differences in prevalence of the different phylogenomic clusters.

Using SLAC, MEME, and FUBAR methods, there was limited detectable diversifying selection for the amino acid substitutions that characterized or defined the different phylogenomic clusters. Only three cluster-defining codons in G (144, 294 and 303) and three in F (125, 172 and 173) glycoproteins were subject to episodic positive selection. It is likely that the observed codon replacements follow non-selective epidemiological processes and these substitutions are compensatory mutations to retain function, or hitchhikers carried along by chance (47), or that immune driven positive selection could not be identified by the three methods used in this study. Nonetheless, the implications of these mutations on protein function, viral evolution and fitness are uncertain and warrant further functional investigation.

A previous study showed that viruses carrying the K68N substitution in the US, affected the binding of MEDI8897 (48). MEDI8897 is an RSV pre-F-specific human mAb with an extended serum half-life, under clinical evaluation as a passive immunization of all infants entering their first RSV season (49). It is probable the K68Q substitution identified in Kilifi promoted evasion of pre-existing immune responses. The unequivocal support for the monophyly of the cluster 20 as an introduction into Kilifi (clade III, **Figure 3(A)**), further supports that this phylogenetic clade was a single (epidemiologically successful) introduction event. Surprisingly, the K68Q F gene amino-acid substitution observed in phylogenomic cluster 20 was not detected as under any selection pressure even though this residue is located at structurally determined mAb binding epitopes (50). Perhaps an explanation for this is the low rate of nonsynonymous evolution (conversely, high sequence conservation) at codon 68 in our dataset. In any case conventional approaches for measuring selection pressure consistently detect positive selection only at codon sites with high rates of nonsynonymous evolution (51).

A high sequence conservation was reported in the MEDI8897 binding site among naturally occurring RSV isolates collected from 1965 to 2014 (50). Our study provides a novel sequence polymorphism (K68Q) within the MEDI8897 binding site with a frequency of nearly 50% in the 2016/17 RSV epidemic in our study population. Functional characterization is required to determine MEDI8897 neutralization and/or binding to viruses containing the K68Q mutation. Additionally, the viruses with the K68Q mutation in 2016/17 epidemic possessed five distinctive amino-acid mutations in G gene, including Y90H and L91F in two consecutive codons. We cannot exclude the possibility that these are relevant antigenic epitopes. Our study underscores the need for continued genomic surveillance of contemporary clinical strains particularly at F and G protein antigenic sites as this has implications on RSV therapeutic and vaccine development.

RSVB viruses containing A103V/L172Q/S173L amino acid-changes in the F protein were also detected during the 2015/16 RSV season in USA and estimated to have likely emerged around 2014 (44) and in China (52), suggesting global circulation of this variant. However, unlike in the US, none of the samples from 2016/17 in Kilifi had these three substitutions probably due to removal by purifying selection.

In conclusion, we present the utility of genomic analyses to investigate virus transmission and genetic diversity including detection of a novel antigenically distinct variant. Further studies are required to determine whether the K68Q mutation is adaptive and/or a result of escape from antibody-mediated selection and constitutes a naturally acquired antiviral resistance-associated mutation that potentially disrupts neutralizing antibody recognition and binding. An important future surveillance effort for us is to assess if the K68Q mutation has become more prevalent and gradually fixed since the 2016/17 epidemic. Additional sequencing of RSVB from other regions in Kenya and neighboring countries is essential to refine evolutionary dynamics and draw better conclusions about geographic origins of viral introductions in the study population in Kilifi. The present study makes publicly available a large number of newly acquired coding-complete RSVB genomes useful for further molecular evolution studies.

## Data Availability

The replication data and analysis scripts for this manuscript are available from the Harvard Dataverse. Genome sequences have been submitted to GenBank.

## Data availability

The replication data and analysis scripts for this manuscript are available from the Harvard Dataverse: ???. Some of the clinical dataset contains potentially identifying information on participants and is stored under restricted access. Requests for access to the restricted dataset should be made to the Data Governance Committee.

## Competing interests

The authors declare no competing interests.

## Acknowledgements

We thank all the study participants for their contribution of samples and data. We also thank the Dispensary / Health Centre management committees for allowing us to conduct the study within their health facilities. We are grateful to the field study team for participant recruitment and the laboratory staff of the KEMRI-Wellcome Trust Research Programme / Virus Epidemiology and Control research group. We would also like to thank D. Collins Owuor for his assistance with the BaTS software and Mark Otiende for his assistance with demographics data. This paper is published with the permission of the Director of KEMRI.

## Funding

This work was supported by the Wellcome Trust [grant 102975, 203077]. CNA is supported by the Initiative to Develop African Research Leaders (IDeAL) through the DELTAS Africa Initiative [DEL-15-003]. The DELTAS Africa Initiative is an independent funding scheme of the African Academy of Sciences (AAS)’s Alliance for Accelerating Excellence in Science in Africa (AESA) and supported by the New Partnership for Africa’s Development Planning and Coordinating Agency (NEPAD Agency) with funding from the Wellcome Trust [107769/Z/10/Z] and the UK government. The views expressed in this publication are those of the authors and not necessarily those of AAS, NEPAD Agency, Wellcome Trust or the UK government.

## Figure legends

**Supplementary Figure 1** (**A**) Violin plots show the distribution (median, IQR) of RSVB qRT-PCR Cycle threshold (*C_t_*) values (<=40) for outpatient participants. (**B**) Comparison of *C_t_* distribution between inpatient (KCH) and the outpatient facilities for children <5 years for the period of December 2015 to June 2017. The threshold used for determining positive and negative samples is shown by dashed line (*C_t_*=35.0).

**Supplementary Figure 2** Frequency (primary Y axis) and proportion (secondary Y axis) of RSV across different age categories in both genders (dashed and dotted lines) from participants presenting to the nine outpatient health facilities over the period December 2015 to June 2017.

**Supplementary Figure 3** Distribution by month of RSVB positive samples collected from the inpatient (severe pneumonia admissions to KCH) and outpatient facilities over the period of December 2015 to June 2017. Secondary Y axis records sample tested (dotted line) and proportion (dashed line) that is RSVB positive per month.

**Supplementary Figure 4** Temporal-scaled phylogeographic maximum clade credibility tree showing sequences of samples collected in Mavueni location (red taxa) in Kilifi County, coastal Kenya.

**Supplementary Figure 5** The number of RSVB cases through time (2015 to 2019) for severe pneumonia pediatric admissions to Kilifi County Hospital. RSV epidemics (usually from October to July of the following year) are indicated.

**Supplementary Figure 6** Maximum likelihood phylogeny of the attachment G glycoprotein gene. The orange colored branches represent the recently emergent variant (phylogenomic cluster 20). The purple colored branches represent the 2018/19 RSVB viruses collected from severe pneumonia pediatric admissions to Kilifi County Hospital.

---

**Supplementary Table 1** Information (accession number, country of sampling and collection date) on RSVB genome sequences of >14,000 nucleotide length retrieved from NCBI GenBank.

**Supplementary Table 2.** Frequency of distribution of symptoms among study participants by 9 outpatient health facilities, age, and gender, between December 2015 and June 2017.

| Characteristic | Fever,<br>% | Chest<br>indrawing,<br>% | Crackles,<br>% | Wheeze,<br>% | Cough,<br>% | Nasal<br>discharge,<br>% | Nasal<br>flare,<br>% | Difficulty<br>breathing,<br>% | Total<br>participants,<br>n |
| --- | --- | --- | --- | --- | --- | --- | --- | --- | --- |
| **Health Facility** |  |  |  |  |  |  |  |  |  |
| Matsangoni | 82.8 | 0.0 | 3.5 | 6.9 | 98.3 | 72.4 | 3.45 | 10.3 | 953 |
| Ngerenya | 84.5 | 1.7 | 1.7 | 0.0 | 98.3 | 65.5 | 0.0 | 10.3 | 898 |
| Soko | 84.0 | 2.0 | 2.0 | 6.0 | 100 | 80.0 | 4.0 | 18.0 | 876 |
| Mtondia | 81.8 | 6.1 | 3.0 | 1.5 | 97.0 | 78.8 | 4.6 | 7.6 | 964 |
| Mavueni | 74.7 | 3.8 | 2.5 | 1.3 | 97.5 | 77.2 | 5.1 | 15.2 | 917 |
| Jaribuni | 79.1 | 2.3 | 4.7 | 9.3 | 97.7 | 83.7 | 0.0 | 4.7 | 846 |
| Chasimba | 65.7 | 3.0 | 1.5 | 6.0 | 97.0 | 71.6 | 3.0 | 9.0 | 893 |
| Junju | 80.0 | 0.0 | 5.0 | 7.5 | 95.0 | 90.0 | 2.5 | 20.0 | 929 |
| Pingilikani | 85.7 | 2.4 | 2.4 | 9.5 | 95.2 | 76.2 | 4.8 | 40.5 | 851 |
| **P value** | **0.156** | **0.588** | **0.974** | **0.142** | **0.900** | **0.192** | **0.741** | **0.001** |  |
| **Age Category** |  |  |  |  |  |  |  |  |  |
| 0–5 mths | 77.7 | 3.5 | 2.4 | 5.9 | 98.8 | 69.4 | 10.6 | 23.5 | 750 |
| 6–11 mths | 83.3 | 8.3 | 4.8 | 6.0 | 100 | 82.1 | 3.6 | 22.6 | 765 |
| 12–23 mths | 84.3 | 2.0 | 3.9 | 5.9 | 97.1 | 73.5 | 1.0 | 13.7 | 1123 |
| 24–35 mths | 82.9 | 1.3 | 2.6 | 4.0 | 96.1 | 81.6 | 2.6 | 9.2 | 788 |
| 3–4 yrs | 79.7 | 0.0 | 1.6 | 6.3 | 98.4 | 76.6 | 1.6 | 9.4 | 962 |
| 5–9 yrs | 83.8 | 0.0 | 2.7 | 2.7 | 91.9 | 83.8 | 0.0 | 8.1 | 1083 |
| 10–19 yrs | 66.7 | 0.0 | 0.0 | 0.0 | 100 | 66.7 | 0.0 | 4.8 | 1045 |
| 20–49 yrs | 52.2 | 0.0 | 0.0 | 0.0 | 91.3 | 87.0 | 0.0 | 4.4 | 1121 |
| 50–100 yrs | 45.5 | 0.0 | 0.0 | 0.0 | 100 | 54.6 | 0.0 | 0.0 | 490 |
| **P value** | **0.003** | **0.041** | **0.883** | **0.830** | **0.129** | **0.150** | **0.009** | **0.010** |  |
| **Sex** |  |  |  |  |  |  |  |  |  |
| Male | 80.5 | 3.5 | 3.0 | 5.6 | 98.7 | 73.2 | 2.2 | 16.5 | 3426 |
| Female | 77.9 | 1.8 | 2.6 | 4.0 | 96.3 | 79.4 | 4.0 | 12.1 | 4700 |
| **P value** | **0.478** | **0.252** | **0.756** | **0.406** | **0.094** | **0.099** | **0.231** | **0.166** |  |

**Supplementary Table 3.** Rate of nucleotide substitution and the estimated date of introduction or emergence of each clade.

| Clade | Estimated date | Rate of evolution ($10^{-3}$ subs/site/y) | | |
| --- | --- | --- | --- | --- |
|  |  | Mean | Lower 95% HPD | Upper 95% HPD |
| I | September 2008 | 1.2 | 0.27 | 2.3 |
| II | July 2007 | 1.18 | 0.33 | 2.32 |
| III | September 2014 | 1.06 | 0.35 | 1.95 |
| IV | August 2012 | 0.43 | 0.26 | 0.54 |
| V | March 2013 | 1.22 | 0.87 | 2.02 |
| VI | August 2012 | 0.67 | 0.58 | 1.1 |
| VII | May 2013 | 1.64 | 1.03 | 2.17 |
| VIII | March 2014 | 0.92 | 0.41 | 1.87 |

**Supplementary Table 4.** Amino acid substitutions identified in RSV genomic regions, which were distinctive to the various phylogenomic clusters.

| Cluster<br>(2015/16) | Amino acid substitutions in genomic<br>regions | Cluster<br>(2016/17) | Amino acid substitutions in genomic<br>regions |
| --- | --- | --- | --- |
| 1 | *P gene* (Ile<sub>60</sub>);<br>*L gene* (Ile<sub>105</sub>, Asn<sub>183</sub>) | 10 | *F gene* (Val<sub>5</sub>, Thr<sub>99</sub>, Thr<sub>129</sub>);<br>*G gene* (Ser<sub>101</sub>, Phe<sub>267</sub>);<br>*NS2 gene* (Asn<sub>53</sub>);<br>*L gene* (Asp<sub>176</sub>) |
| 2 | *N gene* (Val<sub>97</sub>);<br>*G gene* (His<sub>144</sub>) | 11 | *F gene* (Val<sub>5</sub>, Thr<sub>99</sub>, Thr<sub>129</sub>);<br>*L gene* (Asp<sub>176</sub>, Leu<sub>1708</sub>);<br>*G gene* (Phe<sub>267</sub>) |
| 3 | *N gene* (Val<sub>97</sub>) | 12 | *F gene* (Ser<sub>104</sub>, Val<sub>5</sub>,);<br>*G gene* (Phe<sub>267</sub>);<br>*NS2 gene* (Arg<sub>101</sub>);<br>*L gene* (Asp<sub>176</sub>) |
| 4 | *G gene* (Val<sub>290</sub>) | 13 | *F gene* (Val<sub>5</sub>,);<br>*L gene* (Asp<sub>176</sub>);<br>*G gene* (Thr<sub>268</sub>, Ala<sub>290</sub>, Leu<sub>304</sub>) |
| 5 | *NS2 gene* (Thr<sub>5</sub>);<br>*L gene* (Ile<sub>1166</sub>, Arg<sub>2066</sub>);<br>*G gene* (Val<sub>269</sub>, Pro<sub>270</sub>) | 14 | *F gene* (Val<sub>5</sub>,);<br>*L gene* (Asp<sub>176</sub>, Phe<sub>1980</sub>);<br>*G gene* (Thr<sub>268</sub>, Val<sub>290</sub>) |
| 6 | *F gene* (Thr<sub>16</sub>);<br>*L gene* (Ala<sub>1744</sub>, Gly<sub>1787</sub>) | 15 | *N gene* (Val<sub>97</sub>);<br>*G gene* (Thr<sub>205</sub>, Leu<sub>214</sub>, Ala<sub>249</sub>, Leu<sub>293</sub>, Pro<sub>305</sub>) |
| 7 | *F gene* (Leu<sub>172</sub>, Ser<sub>173</sub>, Leu<sub>542</sub>);<br>*G gene* (Ser<sub>75</sub>, Asn<sub>103</sub>, Thr<sub>107</sub>, Arg<sub>136</sub>,<br>Gly<sub>260</sub>, Pro<sub>270</sub>, Ile<sub>279</sub>, Leu<sub>289</sub>, Lys<sub>312</sub>);<br>*M2-1 gene* (Val<sub>181</sub>); | 16 | *F gene* (Cys<sub>25</sub>);<br>*G gene* (His<sub>144</sub>);<br>*L gene* (Glu<sub>304</sub>, Ile<sub>1956</sub>) |
|  | *M2-2 gene* (Asn<sub>26</sub>, Lys<sub>49</sub>);<br>*N gene* (His<sub>216</sub>);<br>*NS1 gene* (Asp<sub>69</sub>, Ile<sub>72</sub>);<br>*NS2 gene* (Thr<sub>26</sub>);<br>*SH gene* (Thr<sub>49</sub>);<br>*L gene* (Leu<sub>56</sub>, Val<sub>715</sub>, Ala<sub>1712</sub>, Asn<sub>1736</sub>,<br>Ile<sub>2019</sub>, Ile<sub>2069</sub>) |  |  |
| 8 | *G gene* (Ser<sub>75</sub>, Ile<sub>76</sub>, Asn<sub>103</sub>, Thr<sub>107</sub>,<br>Leu<sub>110</sub>, Arg<sub>136</sub>, Ser<sub>176</sub>, Leu<sub>191</sub>, Ser<sub>235</sub>,<br>Gly<sub>260</sub>, Ile<sub>279</sub>, Leu<sub>289</sub>, Lys<sub>312</sub>);<br>*F gene* (Ala<sub>103</sub>, Thr<sub>115</sub>, Leu<sub>172</sub>, Ser<sub>173</sub>,<br>Ile<sub>303</sub>, Leu<sub>542</sub>);<br>*M2-1 gene* (Val<sub>181</sub>);<br>*M2-2 gene* (Asn<sub>26</sub>);<br>*N gene* (His<sub>216</sub>);<br>*SH gene* (Thr<sub>32</sub>, Thr<sub>37</sub>, Thr<sub>49</sub>);<br>*L gene* (Leu<sub>56</sub>, Ala<sub>173</sub>, Arg<sub>260</sub>, Val<sub>715</sub>,<br>Ala<sub>1712</sub>, Asn<sub>1736</sub>, Ile<sub>2069</sub>) | 17 | *F gene* (Arg<sub>419</sub>); *NS2 gene* (Thr<sub>5</sub>); *L gene*<br>(Ile<sub>1166</sub>, Arg<sub>2066</sub>); *G gene* (Val<sub>269</sub>, Pro<sub>270</sub>,<br>Asp<sub>294</sub>) |
| 9 | *G gene* (Thr<sub>107</sub>, Arg<sub>136</sub>, Asn<sub>213</sub>, Leu<sub>221</sub>,<br>Ala<sub>227</sub>, Pro<sub>255</sub>, Asn<sub>303</sub>, Ile<sub>279</sub>, Lys<sub>312</sub>);<br>*F gene* (Ala<sub>103</sub>, Pro<sub>125</sub>, Leu<sub>172</sub>, Ser<sub>173</sub>,<br>Thr<sub>19</sub>);<br>*M2-1 gene* (Val<sub>181</sub>);<br>*M2-2 gene* (Asn<sub>26</sub>);<br>*M gene* (Met<sub>73</sub>);<br>*N gene* (His<sub>216</sub>);<br>*SH gene* (Ile<sub>26</sub>, Thr<sub>49</sub>); | 18 | *P gene* (Ile<sub>60</sub>);<br>*L gene* (Ile<sub>105</sub>, His<sub>141</sub>, Asn<sub>183</sub>) |
|  | *L gene* (Leu<sub>56</sub>, Val<sub>715</sub>, Ala<sub>1712</sub>, Asn<sub>1736</sub>, Tyr<sub>1897</sub>, Val<sub>514</sub>) |  |  |
|  |  | 19 | *P gene* (Ile<sub>60</sub>);<br>*L gene* (Ile<sub>105</sub>, Asn<sub>183</sub>);<br>*G gene* (Pro<sub>315</sub>) |
|  |  | 20 | *G gene* (His<sub>90</sub>, Phe<sub>91</sub>, Asn<sub>225</sub>, Ile<sub>273</sub>, Thr<sub>301</sub>);<br>*F gene* (Gln<sub>68</sub>);<br>*NS2 gene* (Arg<sub>80</sub>);<br>*L gene* (Ser<sub>184</sub>) |

## Notes

### Competing Interest Statement

The authors have declared no competing interest.

